# Assessing Cerebral Microvascular Volumetric Pulsatility with High-Resolution 4D CBV MRI at 7T

**DOI:** 10.1101/2024.09.04.24313077

**Authors:** Fanhua Guo, Chenyang Zhao, Qinyang Shou, Ning Jin, Kay Jann, Xingfeng Shao, Danny JJ Wang

**Affiliations:** Laboratory of FMRI Technology (LOFT), Mark & Mary Stevens Neuroimaging and Informatics Institute, Keck School of Medicine, University of Southern California; Siemens Healthcare, USA

**Keywords:** microvascular cerebral blood volume (CBV), vascular space occupancy (VASO), volumetric pulsatility, arterial spin labeling (ASL), ultrahigh-field MRI, age

## Abstract

Arterial pulsation is crucial for promoting fluid circulation and for influencing neuronal activity. Previous studies assessed the pulsatility index based on blood flow velocity pulsatility in relatively large cerebral arteries of human. Here, we introduce a novel method to quantify the volumetric pulsatility of cerebral microvasculature across cortical layers and in white matter (WM), using high-resolution 4D vascular space occupancy (VASO) MRI with simultaneous recording of pulse signals at 7T. Microvascular volumetric pulsatility index (mvPI) and cerebral blood volume (CBV) changes across cardiac cycles are assessed through retrospective sorting of VASO signals into cardiac phases and estimating mean CBV in resting state (CBV0) by arterial spin labeling (ASL) MRI at 7T. Using data from 11 young (28.4±5.8 years) and 7 older (61.3±6.2 years) healthy participants, we investigated the aging effect on mvPI and compared microvascular pulsatility with large arterial pulsatility assessed by 4D-flow MRI. We observed the highest mvPI in the cerebrospinal fluid (CSF) on the cortical surface (0.19±0.06), which decreased towards the cortical layers as well as in larger arteries. In the deep WM, a significantly increased mvPI (p = 0.029) was observed in the older participants compared to younger ones. Additionally, mvPI in deep WM is significantly associated with the velocity pulsatility index (vePI) of large arteries (r = 0.5997, p = 0.0181). We further performed test-retest scans, non-parametric reliability test and simulations to demonstrate the reproducibility and accuracy of our method. To the best of our knowledge, our method offers the first in vivo measurement of microvascular volumetric pulsatility in human brain which has implications for cerebral microvascular health and its relationship research with glymphatic system, aging and neurodegenerative diseases.

## INTRODUCTION

Cerebral arterial pulsation is the rhythmic dilation and constriction of the arteries in the brain with the cardiac cycle. This pulsation results from the pressure wave generated by heart pumping which propagates along the arterial tree. Recent studies have focused on cerebroarterial pulsatility and its potential impact on cerebrovascular and neurodegenerative diseases. It was found that higher arterial pulsatility is generally associated with more severe small vessel disease (SVD)^1–4^ and Alzheimer’s disease (AD) as well as worse cognitive performance^5–8^, highlighting the role of cerebroarterial pulsatility in the pathogenesis and progression of cerebrovascular and neurodegenerative diseases.

Previous studies have attempted to understand the function and underlying mechanisms of cerebroarterial pulsation within the brain. Iliff and colleagues proposed that arterial pulsatility is a key driver of cerebrospinal fluid (CSF) movement in the perivascular spaces (PVS) into and through the brain parenchyma in mice^9^, facilitating the clearance of interstitial solutes and wastes in the brain^10–12^. In contrast, Weller and colleagues speculated that arterial pulsatility provides the main driving force by which interstitial solutes, including amyloid β (Aβ), exit the brain parenchyma along intramural basement membranes of cerebral arteries^13^. In general, arterial pulsation plays a crucial role in driving the glymphatic system^14–18^, a recently discovered macroscopic waste clearance system that utilizes a unique system of perivascular channels within brain. Mestre and colleagues directly observed that hypertension leads to an increase in arterial volumetric pulsatility, which subsequently results in a reduction in CSF flow^19^. This finding underscores the potential influence of alterations in arterial volumetric pulsatility on the glymphatic system and provides a theoretical basis for understanding various cognitive disorders. Additionally, Salameh and colleagues found that arterial pulsation directly influences neuronal activities in the rat olfactory bulb via mechanosensitive ion channels, synchronizing neuronal oscillations with the heartbeat^20^. These studies collectively support the role of arterial pulsatility in promoting fluid circulation in the glymphatic system and even directly influencing neuronal activity. However, all these studies were performed in animal models using invasive optical imaging and electrophysiological recording.

Currently, arterial pulsatility in the human brain is primarily measured using transcranial doppler ultrasonography^21^ (TCD) and phase-contrast MRI^22,23^ (PC-MRI), which were predominantly focused on examining pulsatility in large blood vessels. Recently high resolution (sub-mm) PC-MRI at 7T has been applied for measuring the pulsatility of smaller lenticulostriate arteries^24^, however it remains difficult to assess the pulsatility of cerebral microvasculature including arterioles, capillaries and venules. In addition, existing studies mainly capture blood flow velocity pulsatility, rather than vascular volumetric pulsatility which directly impacts the movement and circulation of surrounding CSF.

Here we proposed a new method to assess volumetric pulsatility of cerebral microvasculature across cortical layers in gray matter (GM) and white matter (WM), using high-resolution 4D vascular space occupancy (VASO) MRI^25^ with simultaneous recording of pulse signals at 7T. Microvascular volumetric pulsatility index (mvPI) and cerebral blood volume (CBV) changes across cardiac cycles are assessed through retrospective sorting of VASO signals into cardiac phases and estimating mean CBV in resting state (CBV_0_) by arterial spin labeling (ASL) MRI^26^ at 7T (refer to Figure 1 and Methods below for details). We applied this method to 11 young and 7 older participants and compared mvPI results and pulsatility of large arteries assessed by 4D-flow PC-MRI. We observed the highest mvPI in the CSF on the cortical surface, which decreased towards cortex as well as in larger arteries. In the deep WM, a significantly increased trend of mvPI was observed in the older group compared to young group. We further performed a series of analysis (such as test-retest scans, non-parametric tests and simulations) to demonstrate the reproducibility and accuracy of our method.

**Figure 1.**
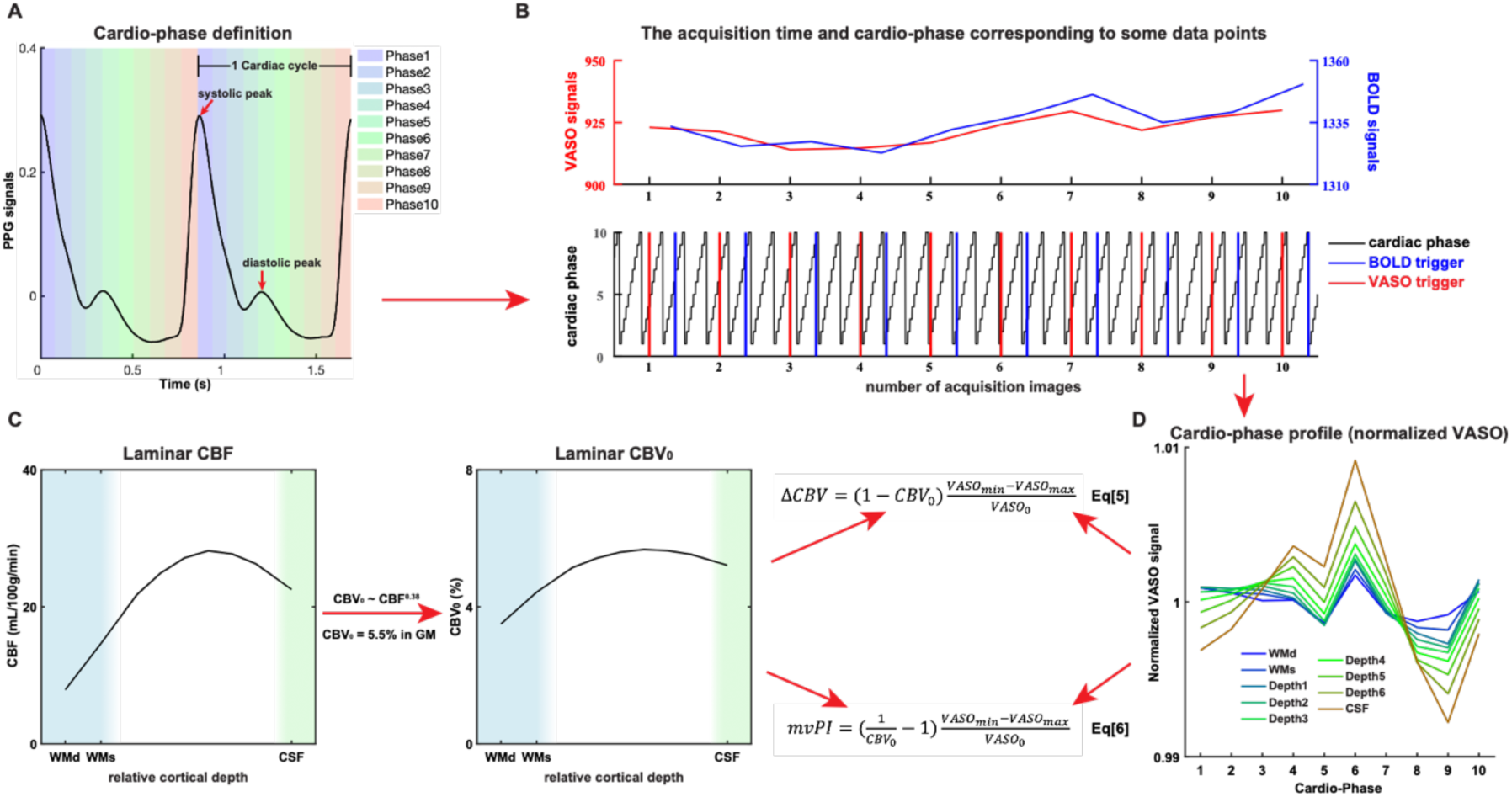
Schematic diagram of microvascular volumetric pulsatility index (mvPI) calculation process. (A) Cardiac phase definition: Two complete cardiac cycles of pulse signals are shown. The time from one cycle’s systolic peak to the next systolic peak is divided into 10 equal phases, defined as phase 1-10. (B) Retrospective sorting of images into 10 cardiac phases: The time and signal intensity curves of VASO (red) and BOLD (blue) signals are shown. The lower figure shows the trigger time corresponding to each image and its corresponding cardiac phase (black). (C) CBF and CBV_0_ laminar profiles based on high resolution (iso-1.25mm) ASL scans and CBV_0_ was derived based on the listed equation. (D) Variations of VASO signals across cardiac phases after retrospective sorting of VASO images, the mvPI was calculated based on the listed equations.

## RESULTS

### Microvascular volumetric pulsatility in CSF, GM, and WM

We first segmented the GM into 6 equi-volume layers (denoted as layers 1-6), as well as one layer of superficial WM and an additional layer of CSF on the cortical surface derived from T1-weighted structural MRI data (Figure S1 shows the schematic diagram of segmentation and estimated cortical depth). A region of interest (ROI) for deep WM was generated by subtracting the ROI of superficial WM from the overall ROI of WM. This allowed us to calculate the laminar profiles of the measured VASO and ASL signals on the cerebral cortex along with their values in cortical CSF, superficial and deep WM respectively (thereafter, refer to deep WM, superficial WM, 6 cortical depth laminae and cortical CSF as cortical laminae). We calculated the laminar profile of CBF from ASL data for each participant. Figure S2A shows a representative cerebral blood flow (CBF) map and Figure S2C shows a zoomed-in view of the superimposed segmentation, demonstrating the high quality of iso-1.25mm CBF maps acquired using our 3D turbo-FLASH (TFL) pseudo-continuous ASL (pCASL) at 7T^26^. Figure 2A illustrates the average laminar CBF and calculated CBV_0_ profiles of 11 young participants. A one-way ANOVA revealed a significant main effect of lamina (F = 12.34, p < 0.001, 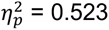 in CBF; F = 131.80, p < 0.001, 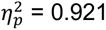 in CBV_0_), indicating differences in resting CBF and CBV_0_ across cortical layers. The CBF profiles exhibit the highest values in the middle and superficial layers of the GM, aligning with the vascular density profiles reported by previous study^27^. Based on Eq 5 in Methods, we estimated the CBV changes across a cardiac cycle (ΔCBV = CBV_max_ − CBV_min_) from VASO signals and plotted the laminar profile of ΔCBV in Figure 2B. A one-way ANOVA revealed a significant main effect of lamina (F = 18.34, p < 0.001, 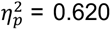), indicating that ΔCBV progressively increases from the WM to superficial layers of the gray matter and CSF. This observation aligns with the reported laminar profile of baseline CBV by Akbari et al^28^. Using the method outlined in Figure 1 (refer to Methods for details), we derived the laminar profile of the mvPI (Figure 2C) in the whole imaging slab. A one-way ANOVA revealed a significant main effect of lamina (F = 12.04, p < 0.001, 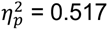), illustrating lamina-specific differences in mvPI. Figure S3A shows a matrix to quantify the differences in mvPI values across various cortical laminae. In conjunction with Figure 2C, we found that mvPI has the highest value in cortical CSF which is significantly higher than all other laminae (Bonferroni correction, factor = 36, all corrected p < 0.001). In general, the cortical CSF exhibits the highest mvPI value (0.19±0.06) and the mvPI gradually decreases as the pial arteries enter the GM, reaching its lowest point in the deep WM.

**Figure 2.**
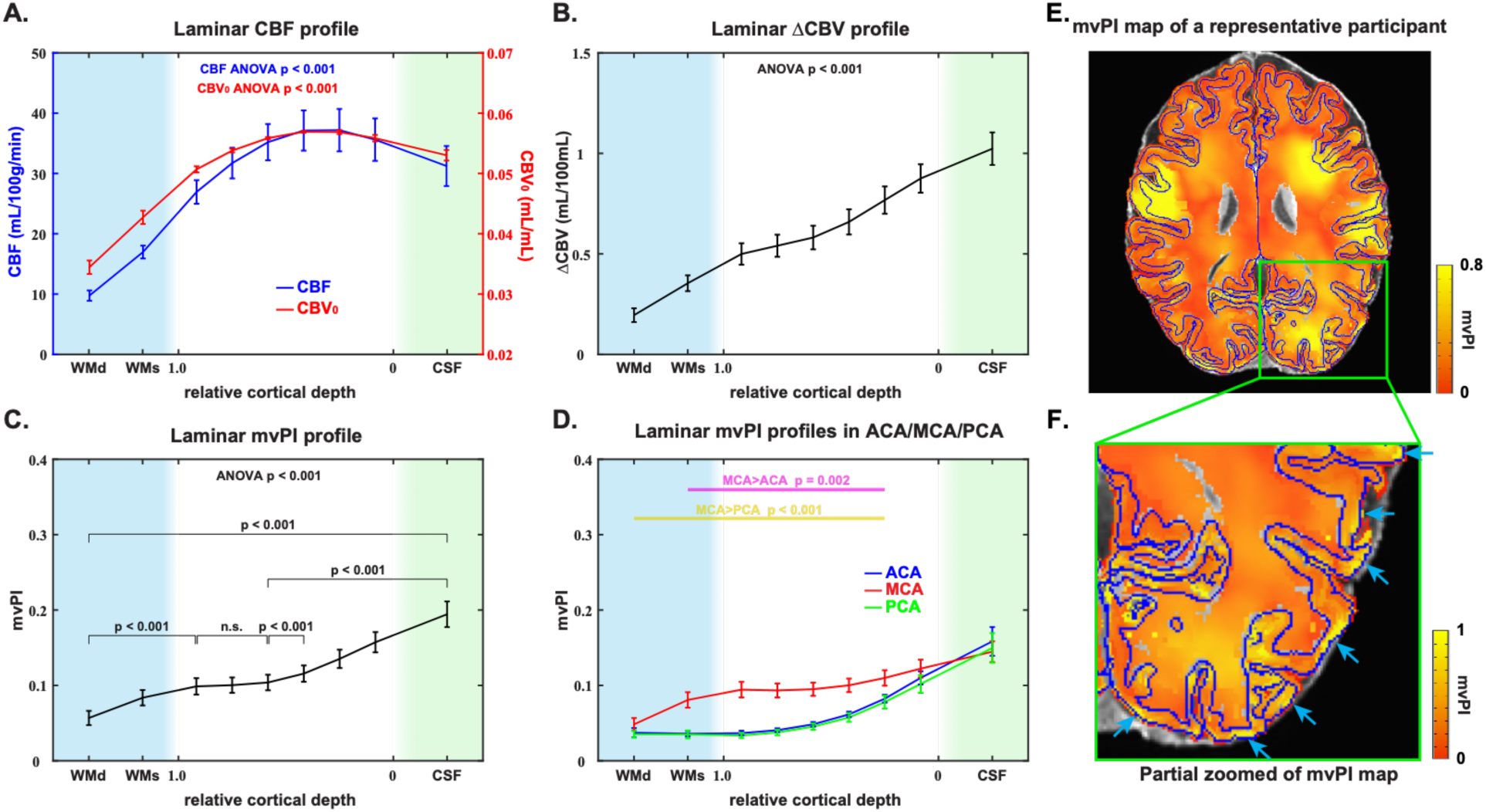
pulsatility maps and laminar results of CBF, CBV_0_, ΔCBV, and mvPI. Laminar profiles of CBF value (A), calculated CBV_0_ (A), ΔCBV (B), and microvascular volumetric pulsatility index (C) show the laminar changes of these parameters. The light blue shaded area indicates the white matter area, and the light green shaded area indicates the CSF area. WMd and WMs in x axis represent the deep white matter and superficial white matter. The values on the x-axis from 1 to 0 represent the gray matter from deep to superficial layers. Bonferroni correction, factor = 36. (D) Laminar mvPI profiles in ACA (blue), MCA (red) and PCA (green) territories. The pink line shows the clustered permutation test result between ACA and MCA. The yellow line shows the clustered permutation test result between ACA and PCA. (E) mvPI maps in a representative participant. (F) Partially zoomed of the mvPI map. The error bar is SEM.

### Microvascular volumetric pulsatility in vascular territories

Given the different vascular compliance^29^ and supratentorial vascular supply region^30^ of the anterior cerebral artery (ACA), middle cerebral artery (MCA), and posterior cerebral artery (PCA), we examined the average mvPI in the 3 major vascular territories. We utilized the vascular territory atlas reported in previous studies^31^ as the ROIs for our analysis. Figure 2D shows the laminar mvPI profiles in ACA, MCA and PCA. A two-way repeated measures ANOVA revealed the significant main effects of lamina (F = 51.84, p < 0.001, 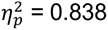) and vascular territories (F = 9.71, p < 0.001, 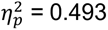) and the significant interaction effect (F = 6.93, p < 0.001, 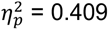), indicating that there are significant differences in mvPI distributions across the 3 vascular territories. Among them, the mvPI in MCA from WM to middle and superficial GM is significantly greater than in ACA (Bonferroni correction, factor = 9, layer 2 corrected p = 0.036, from layer 3 to superficial WM corrected p < 0.001; clustered permutation test, p = 0.002) and PCA (layer 2 uncorrected p = 0.008, from layer 3 to superficial WM corrected p < 0.001, deep WM uncorrected p = 0.006; clustered permutation test, p < 0.001). Overall, cortical CSF has the highest mvPI among all vascular territories. As the pial arteries penetrate deeper into the cortex, the mvPI decreases faster and to a greater degree in ACA and PCA than in MCA.

### Microvascular volumetric pulsatility maps

Voxel-wise mvPI maps were generated with the aforementioned method. Figure 2E displays the mvPI map of a representative participant. It indicates that mvPI is not uniformly distributed across the cortex, with regional high mvPI values observed in the WM. Figure 2F offers a detailed view of the mvPI map in a partially zoomed view of Figure 2E, highlighting the distribution patterns within the GM. The arrows in this figure pinpoint areas with notably high mvPI values located in the superficial GM and CSF. This observation aligns with the finding in the laminar profile, indicating significantly higher mvPI values in the CSF and superficial GM compared to the deep and middle GM. Figure S4A shows the mvPI maps of other participants, demonstrating the high quality of mvPI maps calculated by our proposed method.

### Reliability of mvPI measurements

The results presented above demonstrate a proof-of-concept of our method for mapping mvPI. We then investigated the reliability of our method through three distinct approaches. Firstly, we conducted a test-retest study on three participants with a long interval of 70 days. Secondly, we employed a non-parametric reliability test to evaluate the robustness of our collected data. Lastly, we generated simulated data to analyze the influence of various parameters on the outcomes.

Figure 3A displays the mvPI maps from the two scans of the three participants across different scan sessions. These images reveal that the mvPI distribution patterns remain highly consistent across the two scans for the same individual yet exhibit marked differences across different participants. To accurately quantify the differences between the images, we calculated the mean squared error (MSE) values for each pair within the six maps (more details in Methods). Figure 3B presents the MSE matrix, further strengthening the observed patterns. Figure 3C further visualizes these values, demonstrating a significant distinction in the MSE distribution between maps of the same participant versus those from different participants (p<0.001). This finding underscores the reproducibility of the mvPI maps despite the relatively long interval between the two scans. Moreover, the capability to visualize individual mvPI maps in each participant enhances the potential for future clinical applications.

**Figure 3.**
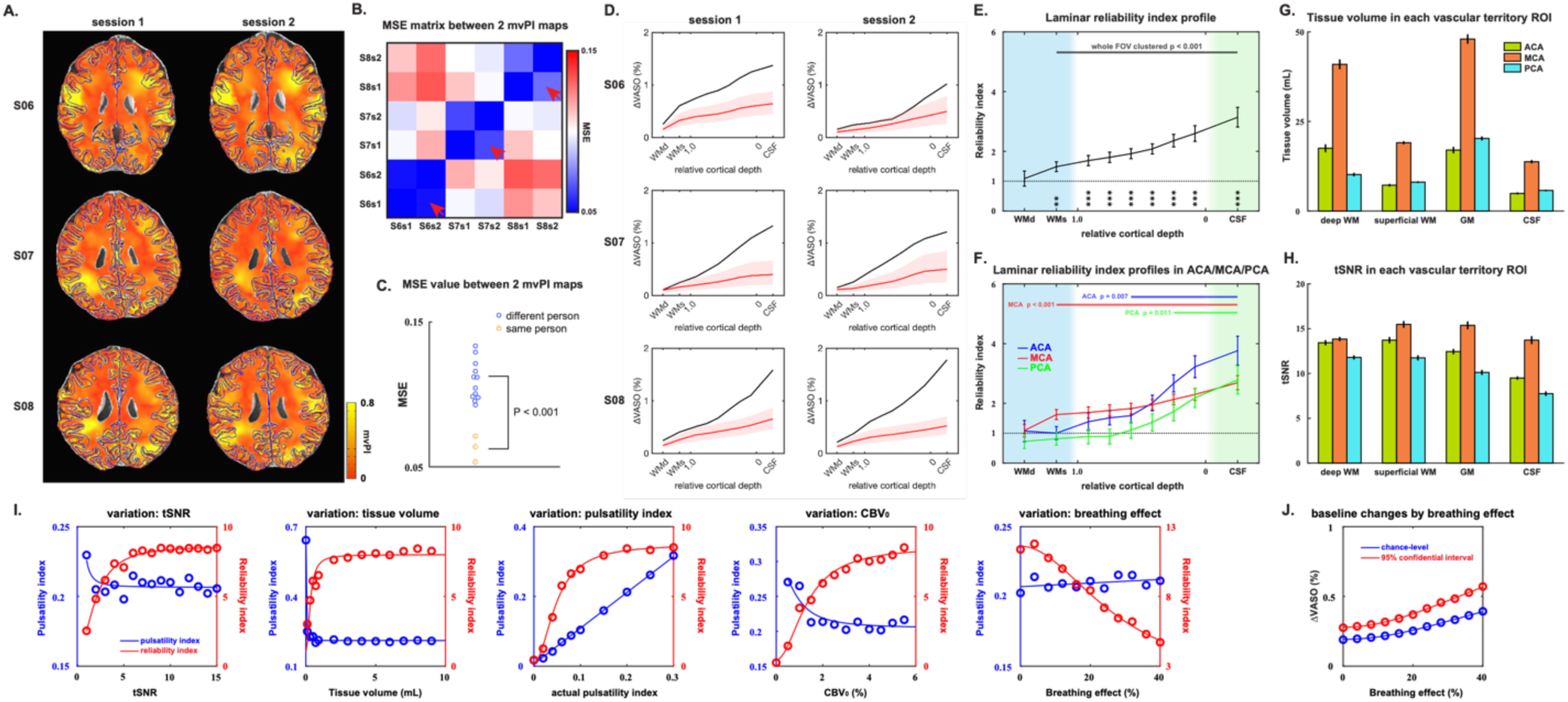
The results of test-retest, reliability test and simulations. (A) mvPI maps in 3 young participants and 2 different scans. (B) MSE value matrix calculated by 6 mvPI maps in figure 3A. (I) Distribution of the MSE values in figure 3B. (D) The results of the reliability test for three young participants with 2 sessions scan data. The red line and red shading represent the mean laminar profile of ΔVASO and its 95% confidence interval, respectively, calculated from random data. The black line shows the laminar profile of the ΔVASO calculated for each subject. (E) Average laminar profile of reliability index (RI) cross the young participants. Bonferroni correction, factor = 9; **: corrected p < 0.01; ***: corrected p < 0.001. The top line shows the clustered permutation test result between RI and chance-level. (F) Laminar RI profiles in ACA (blue), MCA (red) and PCA (green). The top lines show the clustered permutation test results between RI and chance-level in ACA (blue), MCA (red) and PCA (green). The tissue volume (G) and the average tSNR (H) in each vascular territory and each laminar ROI. (I) Results of simulations. It shows the impact of various univariate changes (variations from left to right: tSNR, tissue volume, actual pulsatility index, CBV0, and breathing effect) on the calculated pulsatility index (blue) and reliability index (red). (J) Baseline changes by breathing effect in simulation. It shows the chance-level (blue) and 95% confidential interval upper (red). The error bar is SEM.

As the second proof for the reliability, we employed a non-parametric test by randomly shuffling the time series of the VASO images to artificially create a random signal without synchronization with cardiac phases. A ΔVASO value can be calculated for this batch of data. By repeating this process 10,000 times, we obtained a ΔVASO value distribution from random data (more details in Methods). Figure 3D shows the results of the reliability test for three young participants with test and retest data. The red line and red shading represent the mean laminar profile of ΔVASO and its 95% confidence interval (CI), respectively, calculated from random data. The black line shows the laminar profile of the measured ΔVASO in each subject. When the black points fall above the 95% CI, the VASO signal changes caused by the vascular volumetric pulsatility at this lamina are reliable. Conversely, if the black points fall within the 95% CI, it suggests that the ΔVASO at this depth are indistinguishable from noise, rendering the results unreliable. By comparing the reliability test results of the two scans of the three participants in Figure 3D, it is evident that ΔVASO is reliable at almost all depths and exhibits high repeatability. CSF and superficial GM appear to be more reliable than middle GM, deep GM, and WM. The reliability of ΔVASO in WM is less stable. Figure S5A shows these results of other participants. We also plotted a laminar profile of tSNR of VASO time series (Figure S5C). The distribution of tSNR and chance-level (envelope of 95% CI) show inverse relationship, which illustrates the accuracy of our chance-level calculation. In order to perform inter-group statistical tests, we introduced the reliability index (RI), as shown in Eq 7, which is obtained by dividing the difference between the ΔVASO and the mean of the distribution by the difference between the upper level of the 95% CI and the mean of the distribution (more details in Methods). If the RI value is greater than 1, it means the result is reliable, and vice versa. Figure 3E shows the laminar profile of RI value in the whole imaging slab. Except for the deep white matter (uncorrected p = 0.743), all laminae detected ΔVASO significantly above chance-level (Bonferroni correction, factor = 9, superficial WM corrected p = 0.002, from CSF to layer 6 corrected p < 0.001; clustered permutation test, p < 0.001). It illustrates that the vascular volume changes caused by vascular pulsation in deep WM are too weak to be distinguished from noise, and the mvPI calculated by the other laminae are stable and reliable. Figure 3F shows the laminar profiles of RI value in different vascular territories. It illustrates that the microvascular pulsatility in the ACA (clustered permutation test, p = 0.007) and PCA (clustered permutation test, p = 0.011) could be stably and reliably detected only in the CSF and superficial GM. In contrast, all laminae, except for deep WM, exhibited detectable pulsatility in the MCA (clustered permutation test, p < 0.007). This is consistent with the results that show that the mvPI in MCA is significantly greater than in ACA and PCA. In summary, the results of the reliability test confirm that the mvPI we calculated is genuinely derived from CBV changes induced by vascular pulsation rather than noise. Furthermore, the calculated mvPI exhibits distinct laminar-specific and vascular territory-specific differences.

Considering the numerous factors (such as tSNR, ROI size, breathing effect on vascular volume, CBV_0_, etc.) that may influence the calculated results, we further generated simulated data based on specific parameters from our dataset and conducted an analysis (details in Methods, processing steps shown in Figure S6A). Figure 3I illustrates the impact of various univariate changes (including tSNR of a single voxel, tissue volume, actual volumetric pulsatility index, CBV_0_, and breathing effect) on the calculated mvPI and RI. The simulation results showed that when the voxel tSNR exceeds 5 and other factors remain constant, the calculated mvPI and RI are stable, with the mvPI value closely approximating the set value. In our measurement ROIs, the tSNR generally exceeds 7 (Figure 3H), suggesting that tSNR differences do not significantly impact our results. Additionally, when the tissue volume of an ROI exceeds 2 mL and other factors remain unchanged, the calculated mvPI and RI remain stable, with the mvPI value closely approximating the set value. The tissue volume in our actual ROIs generally exceeds 5 mL (Figure 3G), indicating that ROI size differences do not significantly impact our results. When the univariate change is the actual mvPI, the simulation results demonstrate that we can consistently calculate the mvPI with high accuracy. Additionally, the RI increases with the actual mvPI and reaches a plateau when the actual mvPI approximates 0.2. Given that the maximum mvPI value observed in our results is approximately 0.2 (Figure 2C), a certain degree of correlation can be identified between the RI values we obtained and the mvPI. From the simulation of the effect of CBV_0_ on the results, we observed that changes in CBV_0_ in GM and CSF (approximately >4%) do not significantly affect the calculated mvPI and RI (the fourth plot of Figure 3I and Figure S6C). However, changes in CBV_0_ in WM (approximately 3%~4%) impact the mvPI and RI results (the fourth plot of Figure 3I and Figure S6B). Nonetheless, when the mvPI value exceeds 0.05, the mvPI can always be calculated with high accuracy, and the RI remains stably greater than 1, increasing with the mvPI. This finding is consistent with our results, where the mvPI in deep white matter is approximately 0.05 (Figure 2C), and the RI does not significantly differ from chance level (Figure 3E), indicating that no reliable mvPI value can be obtained. The calculated mvPI remains unaffected by changes in the breathing effect, likely due to the independence of the cardiac and breathing phases. However, the RI decreases significantly with an increase in the breathing effect. This decrease is attributed to the impact of the breathing effect on the chance-level baseline (Figure 3J). Overall, the simulation results support the reliability of calculated results and indicate that genuine differences exist in microvascular pulsatility across different vascular territories and laminae.

### Microvascular volumetric pulsatility index changes with aging

A decreasing CBF, lower metabolic rates of glucose and oxygen and a compromised structural integrity of the cerebral vasculature with special attention to microvasculatures are representative degenerative features of the vascular system of the aging brain^32^. To further explore the application of our new method and to investigate how the distribution and magnitude of mvPI very with aging, we collected datasets from 7 older participants. Following the same analytical procedures, we obtained the laminar mvPI profile for the older participants (Figure 4A) in the whole imaging slab, along with the difference matrix between each lamina (Figure S3B). Compared to the mvPI distribution in young participants (Figure S3C), the mvPI of older participants exhibits a marginally significant increase in deep WM (uncorrected p = 0.029), and the overall distribution pattern is markedly different. The main differences are highlighted by the green circles in Figure S3A and S3B, and by the red statistics in Figure 4A (Bonferroni correction, factor = 36). Figure 4C presents a simplified comparison of the mvPI distribution patterns between the young and older groups. Young participants exhibited a pattern characterized by a monotonic decrease in mvPI from CSF to deep WM. In contrast, older participants displayed a pattern of mvPI decreasing from CSF, reaching a nadir in deep GM, and then remaining constant. Figure 4B displays the distribution of the laminar RI profile in the entire imaging slab for the older group. It demonstrates that the RI in all laminae is significantly greater than 1 (Bonferroni correction, factor = 9, all laminae corrected p < 0.001; clustered permutation test, p < 0.001), indicating that CBV changes induced by vascular pulsation can be stably and reliably detected in all laminae for older participants. In contrast to young participants (Figure 3E), where no reliable CBV changes could be detected in deep WM, older participants exhibited stable and significant CBV changes in deep WM. Additionally, older participants showed significantly higher RI values in both white matter (Bonferroni factor = 9, deep WM corrected p = 0.003 and superficial WM corrected p < 0.001) and deep gray matter (corrected p = 0.008) compared to young participants. Figure S7 presents the laminar mvPI and RI profiles across different vascular territories, demonstrating that, irrespective of the vascular territory, the older participants exhibit higher mvPI and RI values in deep WM supplied by distal vasculature compared to young participants. In conclusion, older participants exhibit higher mvPI and greater reliability in deep white matter compared to young participants. This observation aligns with numerous prior studies that assessed the pulsatility index between age groups, whether based on blood flow velocity in larger arteries of humans^33^ or vascular volumetric changes in rats^19^.

**Figure 4.**
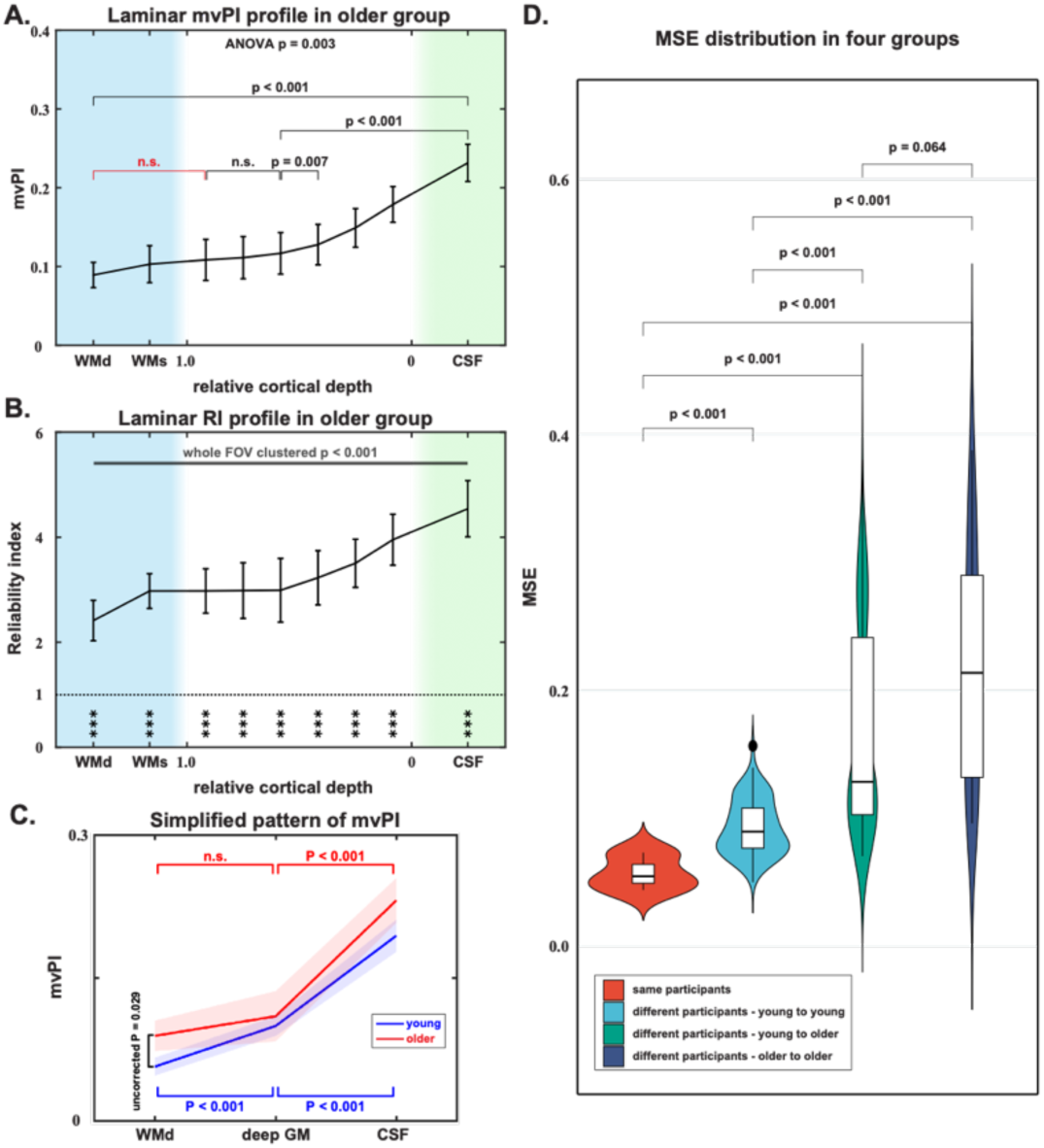
Results of mvPI and RI in older group and the MSE distribution between mvPI maps of different groups. (A) Average laminar profile of mvPI in the older participants. A one-way ANOVA performed a significant main effect of lamina (F = 3.49, p = 0.003, 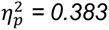). Bonferroni correction, factor = 36. (B) Average laminar profile of RI in the older participants. Bonferroni correction, factor = 9; ***: corrected p < 0.001. The top line shows the clustered permutation test result between RI and chance-level. (C) Simplified pattern of mvPI between young (blue) and older (red). We just chose deep white matter (WMd), deep gray matter (deep GM, average value of deep two points of gray matter) and CSF to compare the mvPI changes along the cerebral microvasculature. (D) MSE distribution in four groups: between two scans of the same participant (red), between different young participants (light blue), between young and older participants (dark green), and between different older participants (dark blue). Bonferroni correction, factor = 6. The error bar is SEM.

Figure S4B displays the mvPI maps of all older participants. Compared to the young (Figure S4A), the mvPI values appear higher in the white matter. This observation aligns with the finding that the laminar profiles of both mvPI and RI values are greater in the older than in the young participants (Figure S3C,D: two-way ANOVAs performed the significant main effect of age p = 0.019 in mvPI and p < 0.001 in RI). Previous studies have suggested that cerebral vascular variability may increase with age^34,35^, and changes in mvPI could contribute to this variability. We employed a similar method to calculate the MSE between all mvPI maps, which provides a comprehensive measure of the differences in mvPI amplitude and distribution between the two maps. We then categorized these MSE values into four groups (as shown in Figure 4D, Bonferroni factor = 6): between two scans of the same participant (group 1, 0.097±0.034, Mean±Std), between different young participants (group 2, 0.186±0.039), between young and older participants (group 3, 0.417±0.256), and between different older participants (group 4, 0.617±0.290). First, MSE in group 1 is significantly lower than the other three groups, indicating that the repeatability of our method remains relatively high. This result further indicates that the uneven distribution of mvPI found in WM is attributable to physiological differences among participants, rather than artifacts or variations introduced by the imaging equipment. MSE in group 2 is significantly lower than group 4, suggesting that the variability of mvPI does indeed increase with age. Finally, standard deviation and mean of MSE in group 3 and group 4 are significantly higher than group 2, demonstrating that the increase in variability is primarily due to the growing differences between older, rather than merely an overall elevation of vPI values in the older participants.

### Pulsatility of velocity and volume in the major cerebral arteries via 4D-flow PC-MRI

Finally, we used 4D-flow PC-MRI to examine the pulsations within the major cerebroarteries and their relationship with the mvPI. Previous studies have demonstrated the feasibility of using 4D-flow to simultaneously measure the pulsatility index of velocity and volume at different locations of blood vessels ^36–38^. As shown in Figure 5A, we selected two ICA sites, one ACA site, three MCA sites, and two PCA sites for pulsatility measurements. Figure 5B shows the cardiac phase profile of velocity and cross-sectional area obtained by ICA for a typical participant (more details in Methods). Similar cardiac phase profiles can be obtained for each ROI. Then according to Eq 8 and Eq 9, we can calculate the pulsatility index of velocity and volume at these locations. Figure 5C shows the velocity pulsatility index (vePI) and volumetric pulsatility index (voPI) calculated for both age groups (young: blue; older: red) within all ROIs.

**Figure 5.**
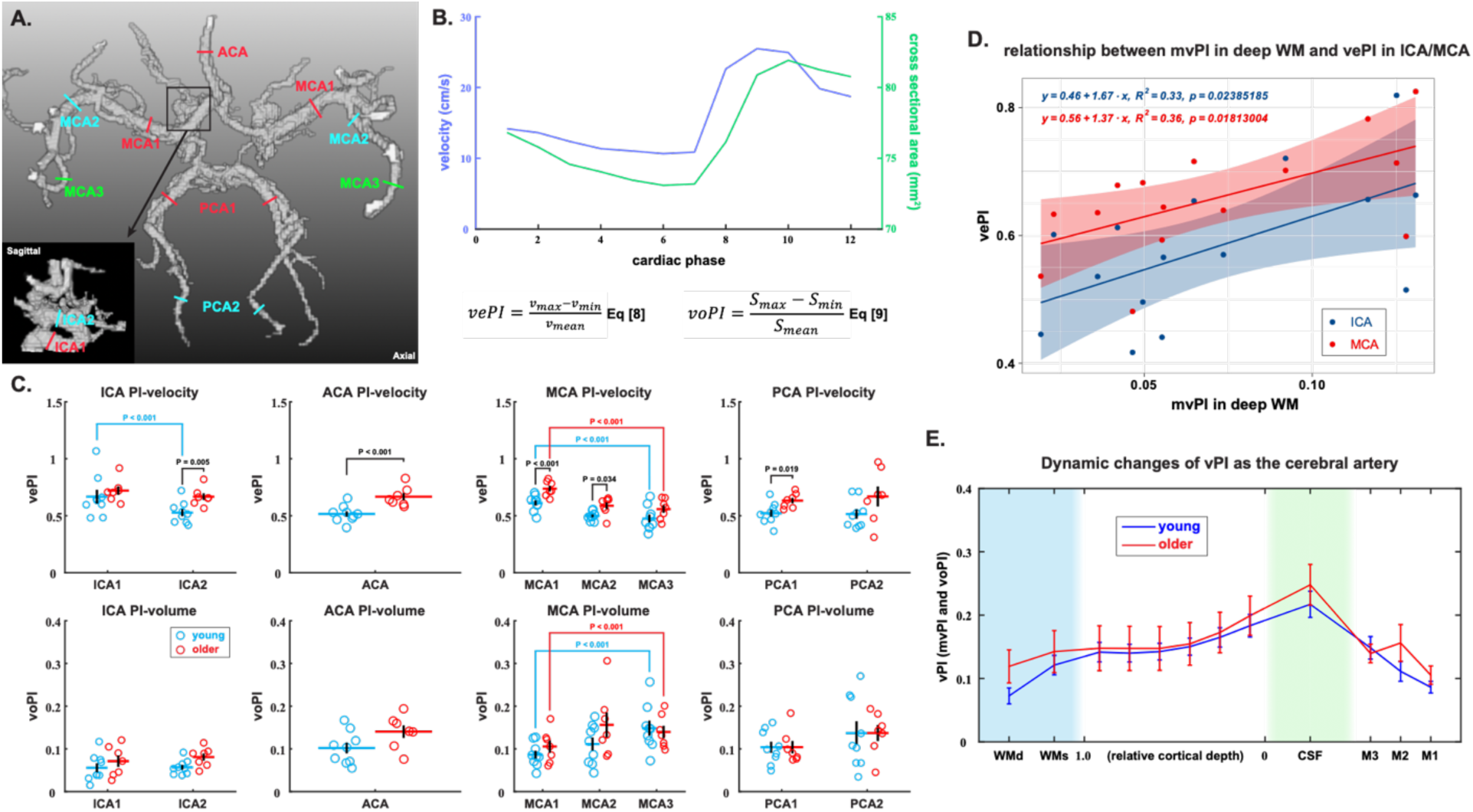
vePI and voPI calculation and results from 4D-flow dataset. (A) Schematic diagram of the ROI selection position of a typical subject in the circle of Willis. The lower left corner is the sagittal plane image within the black frame. (B) The cardiac phase profile of velocity and cross-sectional area obtained by ICA from a random participant. (C) The velocity pulsatility index (vePI) and volumetric pulsatility index (voPI) results in both age groups within all ROIs. The top row of images is the results of vePI, and the bottom row is the results of voPI. From left to right are the results of ICA, ACA, MCA, and PCA. Red is for older subjects and blue is for young subjects. The circles are the results for each participant, and the horizontal line is the average of this group of data. (D) Relationship between mvPI in deep WM and vePI in ICA and MCA1. (E) Changes of volumetric pulsatility index (vPI, including mvPI and voPI) as the cerebral vasculature in MCA territory. The blue and red curves represent the results for young and elder participants, respectively. The light blue shaded area indicates the white matter area, and the light green shaded area indicates the CSF area. WMd and WMs on the x-axis represent the deep white matter and superficial white matter. The values on the x-axis from 1 to 0 represent the gray matter from deep to superficial. M1, M2, and M3 represent the MCA1, MCA2, and MCA3 segment. The error bar is SEM.

Figure 5C displays the mean vePI and voPI calculated for all ROI sites across all subjects. We focused our analysis on the changes in vePI and voPI in relation to arterial blood flow delivery based on the results from the MCA (no significant results were observed in other regions). For vePI, two-way ANOVA (factors: vascular segment and age) showed the significant main effects of vascular segment (F = 18.57, p < 0.001, 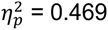) and the vePI in M3 segment was significantly lower than that in M1 (Bonferroni factor = 6, both young and older p < 0.001), indicating that vePI decreases significantly in more distal arteries, align with the previous studies^37,38^. For voPI, two-way ANOVA (factors: vascular segment and age) showed the significant main effects of vascular segment (F = 4.25, p = 0.021, 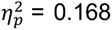) and the voPI in M3 was significantly greater than that in M1 (Bonferroni factor = 6, young group p < 0.001 and older p < 0.001), indicating that voPI increases significantly in more distal arteries, opposing the trend of vePI and has been observed in a previous study^37^. To investigate the effect of age on vePI and voPI, we conducted a two-way ANOVA for vePI which revealed a significant main effect of age (F = 34.54, p < 0.001, 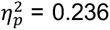), with vePI being significantly higher in the older group compared to the young group in many ROIs (Bonferroni factor = 9, as shown in Figure 5C). This finding indicates that vePI increases with age, consistent with previous studies^33,39,40^ assessing vascular health. In contrast, the two-way ANOVA for voPI showed a marginally significant main effect of age (F = 4.21, p = 0.043, η_p^2^ = 0.036), however no significant difference in voPI was observed between the older and young groups in individual ROIs. Over the past several years, vePI in the ICA and M1 segment of MCA has frequently been used as an indicator to assess vascular health^3,4,36,41^ and vePI values have been shown to be associated with SVD^1–4^ and cognitive impairment in AD^5–8^. Next, we conducted a correlation analysis between the mvPI in deep WM calculated using the proposed method and the vePI in the ICA and M1 calculated using 4D-flow (Figure 5D). Significant correlations exist between the mvPI in deep WM and vePI in the ICA and M1 (Pearson correlation: r = 0.5786, p = 0.0239 with ICA; r = 0.5997, p = 0.0181 with M1).

To characterize the dynamic changes in volumetric pulsatility along the cerebrovascular network, we combined the mvPI results in the MCA territory with voPI measurements from the MCA by 4D-flow (Figure 5E). We found that the volumetric pulsatility index (vPI, including both mvPI and voPI) gradually increased from proximal to distal segments in large arteries, reaching a maximum value in pial arteries within the CSF on cortical surface. Subsequently, the mvPI decreased with increasing cortical depth, reaching a minimum in the deep WM. However, there was a small but significant overall increase in mvPI with aging (2-way ANOVA performed a significant main effect of age, p = 0.019). In particular, there is a marginally significant increase in mvPI with aging within the white matter compared to other locations (uncorrected p = 0.029).

## DISCUSSION

Utilizing our novel method, we obtained the distribution of mvPI in both gray and white matter. Simultaneously, we employed the conventional 4D-flow MRI to determine pulsatility distribution within the large arteries. By integrating these findings, we characterized the dynamic changes in volumetric pulsatility throughout the entire pathway from the large arteries to the arterial terminals in the white matter. We found that arterial volumetric pulsatility peaks in the CSF on the cortical surface, and the vPI gradually decreases in both upstream arteries and downstream microvasculature. Compared with young participants, the mvPI in the gray and white matter of the elderly was overall significantly increased, especially in the white matter.

Cerebral arterioles play a crucial role in the regulating of arterial pressure, resulting in a significant drop compared to large arteries^42,43^. This is primarily due to their smaller diameter, which greatly increases resistance according to fluid dynamics principles (Poiseuille-Hagen formula), where resistance is inversely proportional to the fourth power of the vessel radius. The highly branched network of arterioles further adds to this resistance, causing a marked pressure decrease. As blood flows from larger arteries into arterioles, the total cross-sectional area increases, reducing flow velocity and consequently lowering pressure (as per the continuity equation A_1_×v_1_ = A_2_×v_2_). Arterioles also have smooth muscle cells that actively regulate their diameter through contraction and relaxation, allowing precise control over local blood flow and systemic blood pressure^44,45^. The large arteries cannot significantly reduce arterial and pulse pressures due to their small total volume and large blood vessel diameter^42^. Their primary role is maintaining pulsatile blood flow rather than significantly reducing pressure. In contrast, arterioles, with their higher resistance and regulatory functions, are more effective at dissipating arterial pressure. Therefore, the structure and function of arterioles make them the key sites for arterial pressure reduction in the circulatory system. Arterioles mainly absorb pressure fluctuations through their elastic walls (Figure 6). Expanding and contracting with each heartbeat helps to attenuate the pulse pressure and maintain stable blood flow. The extensive network of small arterioles distributes blood flow and pressure more evenly, significantly reducing the burden on the arterial system and protecting downstream capillaries and tissues from high pressure^42,46^. A cerebral artery on the brain’s surface, including the leptomeningeal and pial arteries, features an endothelium separated from layers of smooth muscle cells by a basement membrane, an intimal layer of extracellular matrix, and often an internal elastic lamina^15,47,48^. As the artery enters the cerebral cortex, it loses its internal elastic lamina and tunica adventitia, becoming an arteriole. The smooth muscle cells in the intima-media of the arteriolar wall on the surface of the cortex are densely arranged, which helps to regulate blood flow and blood pressure. They also contain more elastic fibers and can withstand higher blood flow pressure and dynamic changes^49,50^. This structure enables the arterioles to transport blood from larger arteries to various areas on the surface of the brain and to regulate blood flow to meet the needs of brain activity. In contrast, the number of smooth muscle cells in the arterioles in the cortex is smaller, and their arrangement is not as dense as that of the arterioles on the surface of the cortex^49,50^. They also have fewer elastic fibers and a softer structure. The main function of these arterioles is to supply blood and nutrients to neurons and glial cells inside the gray matter of the brain, participate in local blood flow regulation, and ensure that the metabolic needs of various areas of the brain are met. Arterial blood enters the intracortical arteries through the cortical surface arteries. At this point, the arterial and pulse pressure have been relieved, reducing the pressure that the intracortical arterioles need to release. These position and vascular structure differences result in stronger volumetric pulsatility in the cortical surface arterioles (Figure 6). However, unlike the vPI, the Bernoulli equation establishes a relationship between blood flow velocity and pressure (v^2^ ~ P), indicating that vePI is influenced by both pulse pressure and mean blood flow velocity. As pulse pressure and mean blood flow velocity diminish along the arterial transmission, the vePI correspondingly decreases as well (Figure 6). In summary, from a hydromechanical and hemodynamic perspective, the cortical superficial arteries have the strongest volumetric pulsatility of all arteries in the brain. This may be the reason why we observed that the vascular volumetric pulsatility was strongest in the CSF on the cortical surface.

**Figure 6.**
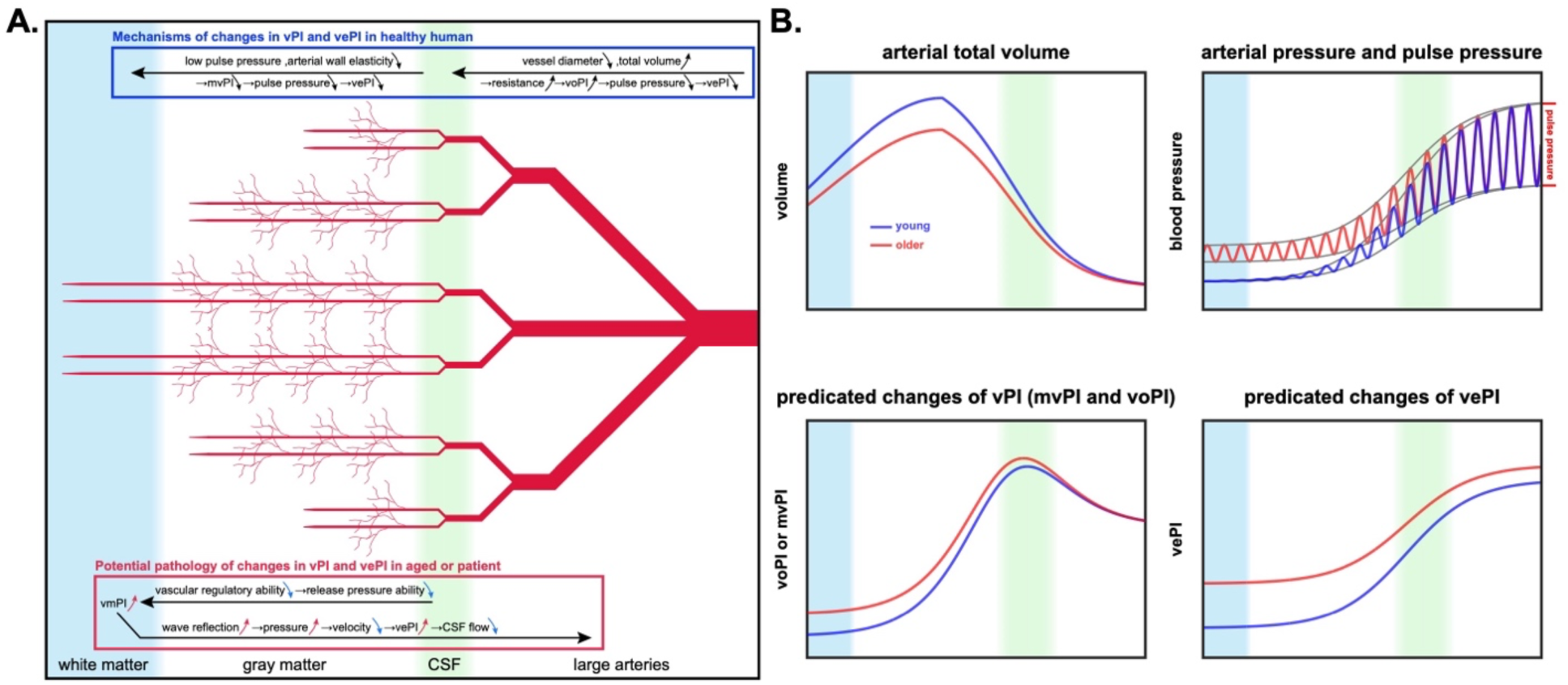
Schematic diagram: physiological mechanisms of laminar changes in arterial pulsatility. (A) Potential physiological mechanisms of arterial pulsatility changes with laminae in normal or abnormal brain. The blue box at the top of the image illustrates the physiological mechanisms underlying the changes in vPI and vePI across arteries in young, healthy brains. Upward-pointing arrows indicate that these parameters increase along the arterial transmission pathway, while downward-pointing arrows signify a decrease. Conversely, the red box at the bottom of the image depicts the mechanisms of abnormal vPI and vePI in elderly or pathological brains. In this context, red arrows indicate an increase in parameters relative to healthy brains, whereas blue arrows denote a decrease. (B) The laminar profiles of arterial total volume, blood pressure, predicated volumetric pulsatility index (including mvPI and voPI) and vePI.

In addition to alleviating pulse pressure and protecting the small blood vessels and capillaries of the brain, the volumetric pulsation of the cerebral arteries has also been found to be a main driver in promoting fluid circulation and waste removal^9,13,15,51,52^. Injection of molecules with widely different molecular weights into the caudate nucleus of rats and CSF of cats revealed that interstitial solutes and CSF use the perivascular space (PVS) for drainage and that molecules with widely different molecular weights have the same half-life, suggesting that fluid exchange is a large volume flow of fluid along this channel rather than diffusion^10,53–55^. Louveau and colleagues proposed that these perivascular spaces serve a pseudolymphatic function, removing metabolic waste and delivering fresh CSF to the interstitium^16^. Of particular importance is that the cyclical changes of the blood vessel wall with the arterial pulsation are the driving force behind the movement of CSF^19^. In summary, the strong volumetric pulsation of the arteries on the cortical surface, discovered by our novel method, can push a large amount of fresh CSF into the brain parenchyma, thereby promoting fluid circulation and waste removal.

Aging brains exhibit several degenerative changes in the vascular system, including decreased cerebral blood flow (CBF), lower metabolic rates of glucose and oxygen, and compromised structural integrity, particularly in microvessels^32^. The reduction in CBF with aging likely reflects a shift in vasoregulatory capacity toward constrictor responses, manifesting as impaired vasodilation or enhanced vasoconstriction^56–58^. Additionally, age-related impairment in perivascular neural innervation, affecting the cerebrovascular regulatory circuit, contributes to lower perfusion rates in the aging brain^59,60^. Numerous studies have reported a reduction in cerebral microvasculature density with aging in both humans^61^ and experimental animals^62–64^. Typical vascular anomalies in aging include perivascular collagen deposits (also known as microvascular fibrosis) and basement membrane thickening (BMT)^65–68^. These factors reduce the total volume of arterioles and their branching, diminishing their capacity to dissipate arterial and pulse pressures^69^. The unreleased pressures are transmitted backward along the arterioles until they reach the distal arteries in the white matter, the end of the arteries. The arterioles in the white matter cannot release these abnormal pressures, leading to a mechanical increase in volumetric pulsation and the generation of strong wave reflections^58,70^. The effective reflection distance of the reflected wave increases in the elderly compared with the young^70^, indicating that the main reflection points of the reflected wave move distally (i.e., the main source of wave reflection changed from various arterial branches to the reflection of the white matter artery at the end of the artery) and support our inference according to fluid mechanics. This strong wave reflection and the superposition of pulsating blood is a complex process, and we cannot yet infer whether it will enhance or weaken vascular volumetric pulsation. However, through the results of this study and previous study on rats^19^, it is inferred that this process ultimately leads to the enhancement of arterial vascular volumetric pulsation and the enhancement is greater the closer to the arterial terminal. However, because the changes in vascular volume caused by this process are influenced not only by arterial blood directly from the heart but also by wave reflections, the time-change curve of the vascular wall is altered. This ultimately disrupts the synchrony between the driving force of vascular pulsation and CSF flow, leading to partial reflux of CSF, significantly slowing down the CSF flow rate^19^. In summary, with aging, the final change pattern is that the volume pulsation of the white matter arteries increases significantly while the volume pulsation of the other arteries increases slightly (Figure 6).

In addition to aging, many disease factors, such as hypertension and arteriosclerosis, can also cause abnormal increases in arterial volumetric pulsation^69,71^. Over time, blood vessels subjected to prolonged higher-than-normal volumetric pulsation are at an increased risk of degeneration (such as vascular fibrosis), especially the small blood vessels in the white matter^72^. This heightened risk can lead to conditions such as small vessel disease (SVD) and cerebral microbleeding^72–74^. If this process continues, there is a higher risk of developing stroke, dementia, cognitive impairment, and other related conditions^73,74^. On the other hand, high arterial volume pulsatility slows down CSF flow^19^ and waste removal. Over time, the accumulation of metabolic wastes that cannot be effectively removed leads to large molecular clusters, such as beta-amyloid protein deposits, further hindering waste removal. This may eventually lead to a series of related diseases such as Alzheimer’s disease^73–75^. Therefore, detailed information about vascular volume and pulsation can help evaluate vascular diseases such as small vessel disease and cognitive diseases such as Alzheimer’s disease and can evaluate the causes of the disease from more aspects.

The reliability of our method is strongly supported by rigorous validation processes. We employed non-parametric testing to confirm that our results are significantly higher than chance-level distributions, ensuring they stem from genuine volumetric pulsatility changes rather than noise. Consistency across multiple scans of the same participants further demonstrates the stability of our method, with similar mvPI distribution patterns observed despite relatively long time gaps. Moreover, we compared the differences between mvPI maps of different participants, finding significantly lower variability within individual participants’ scans than between different individuals. We observed an uneven distribution of mvPI in white matter, with areas of high mvPI showing a strong resemblance to the probability density maps of white matter hyperintensity (WMH) reported in previous studies^76,77^. This suggests that microvascular volumetric pulsatility may be associated with the development of WMH. This high reproducibility and stability underscore the robustness of our approach. The strong correlation between the mvPI results by our new method and the vePI results obtained using 4D-flow further validates the reliability of the new method. Additionally, our findings align with previous in vivo studies, particularly the high mvPI in CSF^9,15,16,19,51^ and significantly increased mvPI in white matter microvasculature with aging in animal models^19^. These factors collectively affirm the reliability and potential of our method as a valuable imaging tool for assessing microvascular health in a living human brain. However, our method has a limitation in that we cannot determine a specific CBV_0_ value for each participant and must rely on an approximate value derived from previous literature. We attempted to set a different CBV_0_ value (0.04) and found that the results remained largely consistent with those of the current study (Figure S8). While the calculated mvPI values differed, the statistical analysis and distribution patterns were essentially unchanged.

Our new method shows great potential in both research and clinical applications. Combining high-resolution VASO and 3D TFL-pCASL MRI, we can accurately image the cerebral microvascular system, capturing volumetric pulsatility in gray and white matter. This approach supports the study of neurodegenerative diseases and the cerebral glymphatic system, with significant implications for early diagnosis, monitoring and exploring basic mechanisms. Overall, our method opens new avenues for research and clinical applications on microvascular pulsatility. However, a limitation of our method is that it relies on an estimated baseline cerebral blood volume (CBV_0_) value rather than determining it specifically for each participant. Although this approximation did not significantly affect our overall findings, it introduces some uncertainty in the exact mvPI values. Further validate of our method in clinical population is warranted.

In summary, to the best of our knowledge, our method combining high-resolution VASO and 3D TFL-pCASL MRI offers the first in vivo measurement of microvascular volumetric pulsatility in the human brain. Our findings of the highest mvPI in the CSF on the cortical surface and a significantly increased mvPI in the WM of older participants compared to younger ones have implications for cerebral microvascular health and its changes with aging and neurodegenerative diseases. Our technology offers a valuable imaging tool for understanding hydromechanical effects on cerebral microvasculature, fluid circulation and brain health.

## Data Availability

All data produced in the present study are available upon reasonable request to the authors

## ACKNOWLEDGEMENTS

The authors thank Dr. Laurentius Huber for sharing the VASO sequence and thank Dr. Junhao Wen for guiding the writing. This work was supported by US National Institute of Health (NIH) grant UF1-NS100614, S10-OD025312, R01-NS114382, R01-EB032169, RF1AG084072, R01-EB028297, R01-NS134712 and R01-NS121040.

## METHODS

### Participants

11 young volunteers (mean age = 28.4 years, SD = 5.8 years, 5 females) and 7 older volunteers (mean age = 61.3 years, SD = 6.2 years, 4 females) participated in this study. All participants in this study were healthy without known neurological or psychiatric disorders and refrained from caffeine 3 hours before the scan. All participants provided written informed consents according to a protocol approved by the Institutional Review Board (IRB) of the University of Southern California. Head motion was minimized by placing cushions on top and two sides of head. All participants underwent pCASL and VASO scans with recording photoplethysmogram (PPG) signals on the finger by BioPac. Three of the young participants (S06, S07, and S08) underwent repeated VASO scans on 2 different days (~70 days interval) for test and re-test. 9 of the young participants and all the older participants were scanned with a 4D-flow sequence.

### MRI data acquisition

MRI data were acquired on the investigational pTx part of a 7T Terra system (Siemens Medical Systems, Erlangen, Germany) with an 8Tx/32Rx head coil (Nova Medical, Cambridge, MA, USA). Anatomical images were acquired with a T1-weighted MP2RAGE sequence (0.7-mm isotropic voxels, 224 sagittal slices, FOV = 224×224 mm^2^, TR = 4500 ms, TE = 3.43 ms, TI1 = 1000 ms, flip angle = 4°, TI2 = 3200 ms, flip angle = 5°, bandwidth = 200 Hz/Px, slice partial Fourier = 6/8, GRAPPA = 3).

Functional images were acquired for resting states with a slice-selective slab-inversion (SS-SI) VASO^25,78–80^ sequence (1-mm isotropic voxels, 24 axial slices, TI1/TI2/shotTR/TE = 1367.4/2512.2/47.7/16.2 ms, partial Fourier factor = 6/8, variable flip angle scheme with reference flip angle = 33°, 3-fold GRAPPA acceleration in z-direction with a CAIPIRINHA shift of 1, bandwidth = 1644 Hz/Px, FOV = 160 mm, matrix size = 160 × 160, acquisition time ~30 min), a 3D TFL-pCASL^26,81^ sequence (1.25-mm isotropic voxels, 80 axial slices, FOV = 224 × 200 × 100 mm^3^, flip angle = 8°, TE = 2.5 ms, TR = 7 s, 10 segments, Poisson-disc undersampling with R = 4, acquisition time ~12 min, Compressed Sensing reconstruction) and a 4D-flow PC-MRI sequence (0.8-mm isotropic voxels, 40 axial slices, FOV = 200 × 200 × 32 mm^3^, reconstructed cardiac phases: 12, flip angle = 15°, TR/TE = 64.32/2.87 ms, SENSE: 3, RL, AP and FH velocity encoding: 100 cm/s, acquisition time ~8 min, Compressed Sensing reconstruction).

### MRI data preprocessing

MRI data were analyzed using AFNI^82^, FreeSurfer^83^, ANTs^84^, SPM12^85^, the mripy package (https://github.com/herrlich10/mripy) and MATLAB (R2021a). The preprocessing of the VASO data includes removing spikes in time series, linear motion correction, and T1w anatomical image registration to functional volumes. ASL data were reconstructed and CBF values were calculated using the method in previous studies^26,81^. The preprocessing of the 4D-flow data included motion correction, phase unwrapping and region of interest (ROI) selection. We estimated a 12-parameter linear transformation from the anatomical volume to a MNI space template by nonlinear transformation with ANTs. Then we can get a vascular territory mask map from the atlas^31^.

### Cortical layers definition

VASO and ASL images were separately upsampled to a finer grid of 0.5 mm^3^ and 0.625 mm^3^ isotropic spatial resolution to avoid singularities at the edges in angular voxel space using the ‘3dresample’ program in AFNI with the Nearest-Neighbor (NN) interpolation method. The T1w MP2RAGE anatomical volume was segmented into white matter (WM), gray matter (GM), and cerebrospinal fluid (CSF) using FreeSurfer’s automated procedure and its high-resolution option. A boundary-based algorithm was separately used to co-register structural MP2RAGE images to VASO and ASL control images^86^. Eight cortical layers (1 WM, 6 GM, and 1 CSF) were calculated using AFNI/SUMA and custom python codes in mripy package with the equi-volume layering approach^87^. Although the cortical depths of different brain regions vary (From ~4mm thickness of the prefrontal cortex to ~2mm thickness of the primary sensory cortex) and the voxel size is not large enough to clearly distinguish the cortical layers, a large number of voxels randomly sampled at different cortical depth allowed us to derive a reliable continuous laminar profile of neural activity^25^, as shown in Figure 1S.

### Pulsatility index calculation in VASO

Previous studies generally used (V_max_-V_min_)/V_mean_ (V is velocity or blood flow) as the pulsatility index calculation equation^36,88^. To estimate the volume change of microvasculature across the cardiac cycle, we used the following formula to calculate the microvascular volumetric pulsatility index (mvPI):

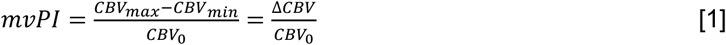

where CBV_max_, CBV_min_ and CBV_0_ are the CBV values when the blood vessels expand to their maximum volume, contract to their minimum volume and averaged over the entire cardiac cycle respectively. ΔCBV is absolutely CBV changes from CBV_min_ to CBV_max_. According to the principle of VASO sequence^89^:

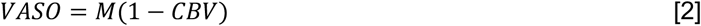

Eq 2 can be converted into the following:

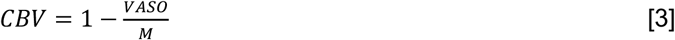

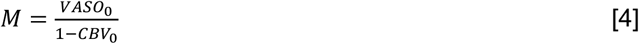

According to Eq 3 and Eq 4, we can deduce:

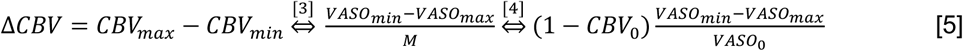

where VASO_min_ is the VASO signal when it reaches CBV_min_, VASO_max_ is the VASO signal when it reaches CBV_max_. Substituting Eq5 into Eq1, we can derive the equation for mvPI:

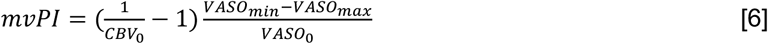

Figure 1 shows the calculation process of the mvPI using VASO and ASL data, including: (1) According to the recorded PPG signal, we defined 10 cardiac phases as shown in Figure 1A. Then we can get the cardiac phase at any time point like this black line in Figure 1B bottom; (2) Since the SS-SI VASO^25,78–80^ sequence recorded both VASO and BOLD image series, we recorded the trigger time of all VASO images and BOLD images (blue and red bar in figure 1B bottom). According to the cardiac phase corresponding to the trigger time, we can define the cardiac phase of every VASO/BOLD image (Figure 1B top); (3) We rebinned all the VASO and BOLD images with same cardiac phase and averaged it. Then we got the cardiac phase profiles of both VASO and BOLD images; (4) We divided the VASO signal by the BOLD signal to obtain the cardiac phase profile of the corrected VASO signal (Figure 1D). From this, we can calculate the value of (VASO_min_-VASO_max_)/VASO_0_ term in Eq 6; (5) According to the ASL results, we can calculate the laminar profile of CBF value (figure 1C left). (6) We assumed CBV_0_~CBF^0.38^ according to the Grubb’s study^90^. Assuming average baseline CBV_0_ = 0.055 ml/ml in the gray matter^91^, we can get the laminar profile of CBV_0_ value (figure 1C right). This is the 1/CBV_0_-1 term in Eq 6. Finally, we can calculate the mvPI value.

Voxel-wise mvPI maps were generated with the aforementioned method. To enhance the signal-to-noise ratio, which may be insufficient in a single voxel, we applied intra-layer spatial smoothing with a 6 mm full width at half maximum (FWHM) prior to calculation^25,92^. Due to the differing voxel sizes between the ASL and VASO sequences, as well as the relative low signal-to-noise ratio of individual ASL voxels, we utilized an average CBF value within each laminar ROI to estimate mvPI.

### Reliability test

We verified the reliability of the results using a non-parametric test. We selected an ROI to extract the average corrected VASO time series, and the ΔVASO of this ROI with cardiac cycle can be calculated using the calculation method described above step 1-4. Then we randomly shuffled the order of this time series and performed the calculation again using the same method to get a ΔVASO value. Repeating this step 10,000 times, we can get a ΔVASO value distribution of results representing no vascular pulsatility with a sample size of 10,000. We can get the mean and 95% confidence interval (CI) of this distribution, and we can also get the P value that our result satisfies the non-hypothesis that the measurement belongs to this distribution (the number of results in the distribution that are greater than the original ΔVASO divided by the sample size). We further defined a reliability index (RI) according to the following equation:

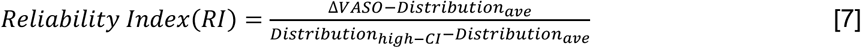

where ΔVSAO is the calculated result from the original time series, Distribution_ave_ is the average value of the random distribution, Distribution_high-CI_ is the upper bound of the 95% CI. As shown in Figure S5B, all participants showed reliable measurements (RI > 1) except for S15. Therefore, we used 6 older participants for mvPI analysis. The RI provided an objective and quantitative metric for quality control of our mvPI results.

### Pulsatility index calculation by 4D-flow

We drew several spherical masks with diameters larger than the diameter of the blood vessels at the locations of straight segments of major blood vessels. These include 2 ICA, 1 ACA, 3 MCAs (M1, M2, and M3 segment) and 2 PCAs (P1 segment and P2 segment), shown in Figure 5A. We set the center of the spherical mask at the center of the blood vessel. The signal intensity of the magnitude image in that region was normalized by the average values of the highest 5% voxels in the mask. Select the 10 voxels closest to the center of the mask and inside the blood vessel to get the blood flow direction. A plane perpendicular to the blood flow direction and passing through the center of the mask can then be obtained, and all voxels with a vertical distance less than 0.5 mm from this plane are selected as the ROI mask at this position. For more fine-grained calculations, we interpolated the voxel to 0.4 mm isotropic for subsequent calculations. For every cardiac phase, we selected voxels that are greater than 20% of the maximum value of the magnitude image and whose blood flow velocity changes with the cardiac cycle by more than 10% as the vascular mask in the ROI. We can get the cardiac phase profiles of blood flow velocity and volume like Figure 5B. Then we calculated the PI values using the following equations:

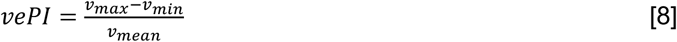

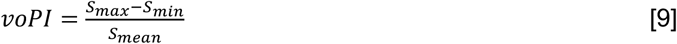

where v means blood flow velocity and S means vessel cross-sectional area. vePI and voPI present the PI values of the changes in blood flow velocity and arterial volume respectively.

### Calculation of mean squared error (MSE)

Image stability assessment such as mean squared error (MSE)^93^ and PSNR^94^ are widely used to measure the quality and stability of image processing. We projected all vmPI maps into MNI space using the nonlinear projection parameters calculated by ANTs. We used the common mask intersection of all maps as the new mask, so all calculations used only the same batch of voxels (for Figure 3B,C). Then we calculated the MSE value between each pair of maps using the following formula:

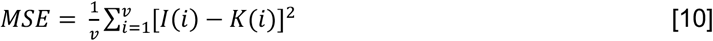

where I and K are two maps, and v is the number of voxels in the mask.

We intended to perform this analysis on all participants. However, due to the small number of remaining voxels from the intersection mask of all participants, we calculated the MSE between two mvPI maps using the intersection of these two mvPI maps instead of the intersection of all masks (for Figure 4D).

### Simulations

Many previous studies have shown that physiological factors such as cardiac cycle and respiratory cycle can affect CBV and fMRI signals^95–97^. Figure S6A displays the simulation process. We assumed that the factors that affect CBV changes are mainly the cardiac phase and respiratory phase and generated CBV time series using sine function. The heart rate was set to 80±10 (mean ± std) times/min and the respiratory rate was set to 12±3 times/min. The pulsatility index and respiration index were both set to 0.2 (based on the results in Figure 2). In this way, we can obtain the time series of CBV changes with the cardiac phase and respiratory phase, respectively. Then, according to the repeated acquisition time of our sequence (about 3.1s), we resampled and obtained the time series of CBV changes with a length of 600 TRs. We added these two time series together and calculated the time series of the VASO signal without noise according to Equation 2. We then added gaussian noise to the time series of the VASO signal to make the tSNR of a single voxel equal to 7 (the smallest tSNR in Figure 3H). We set the ROI size to 5 mL (i.e., 5000 voxels, the minimum value in figure 3G), and then used the proposed method to calculate the pulsatility index and reliability index of the generated data. Finally, we can control a single variable to generate a curve showing the effect of a single variable change on PI and RI values, shown in Figure 3I,J.

### Statistical analysis

The statistical tests in this study were performed using MATLAB and JASP software (version 0.16.4). Given the relatively small sample size of our study, we used non-parametric bootstrapping method for the hypothesis tests of cortical laminae-dependent effect and age effect. Cluster-based permutation test was used for the clustered significance of a range of cortical laminae in vmPI and RI. One-way ANOVA and two-way ANOVA were performed for laminar effect or interaction effect of laminae and age or vascular territory.

## SUPPLEMENT

**Figure S1.**
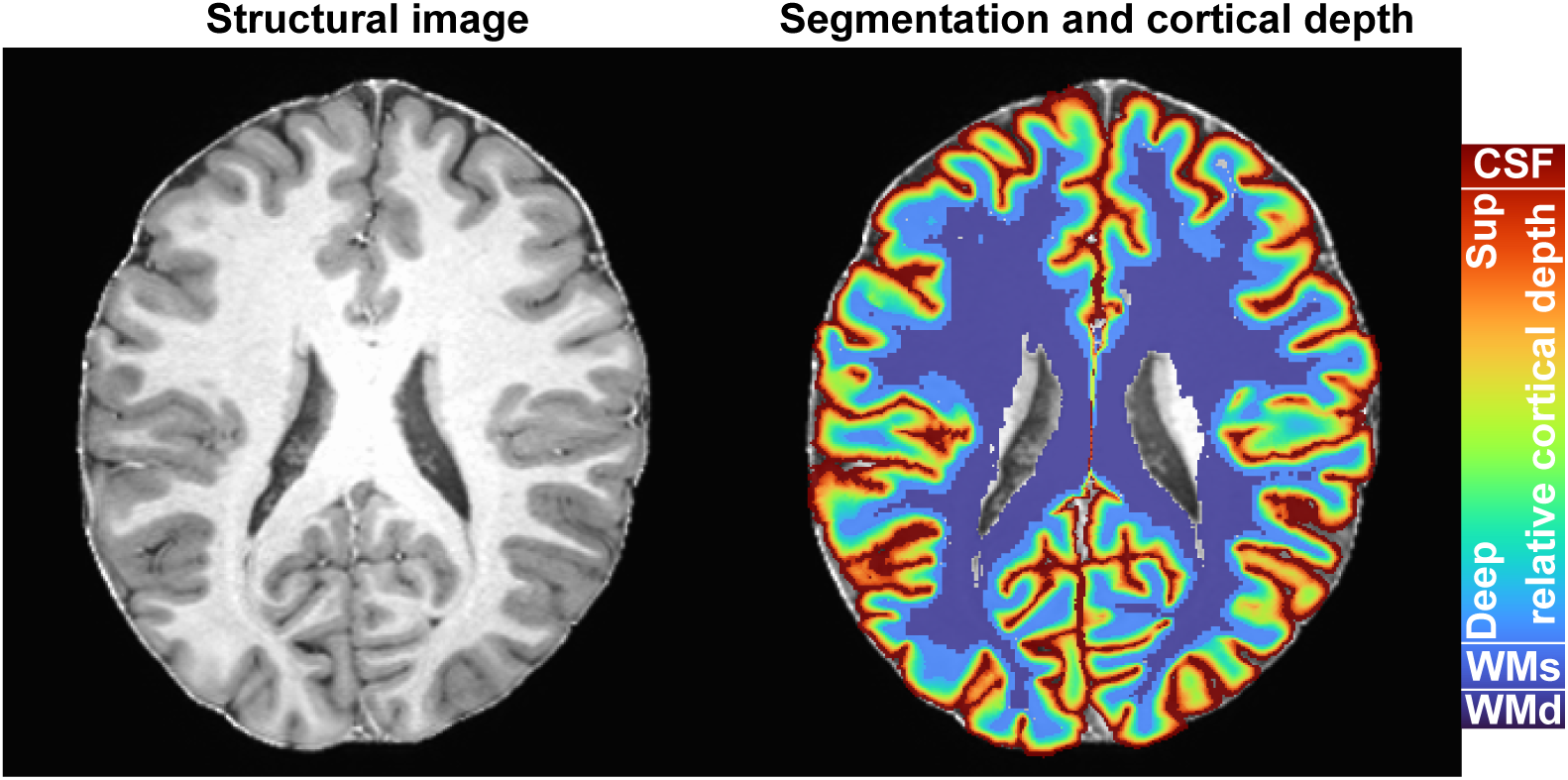
Schematic diagram of segmentation and estimated cortical depth. Left image is the anatomical image from T1w MP2RAGE sequence. Right image is the results of the segmentation and estimated cortical depth. WMd: deep white matter; WMs: superficial white matter; CSF: cerebrospinal fluid.

**Figure S2.**
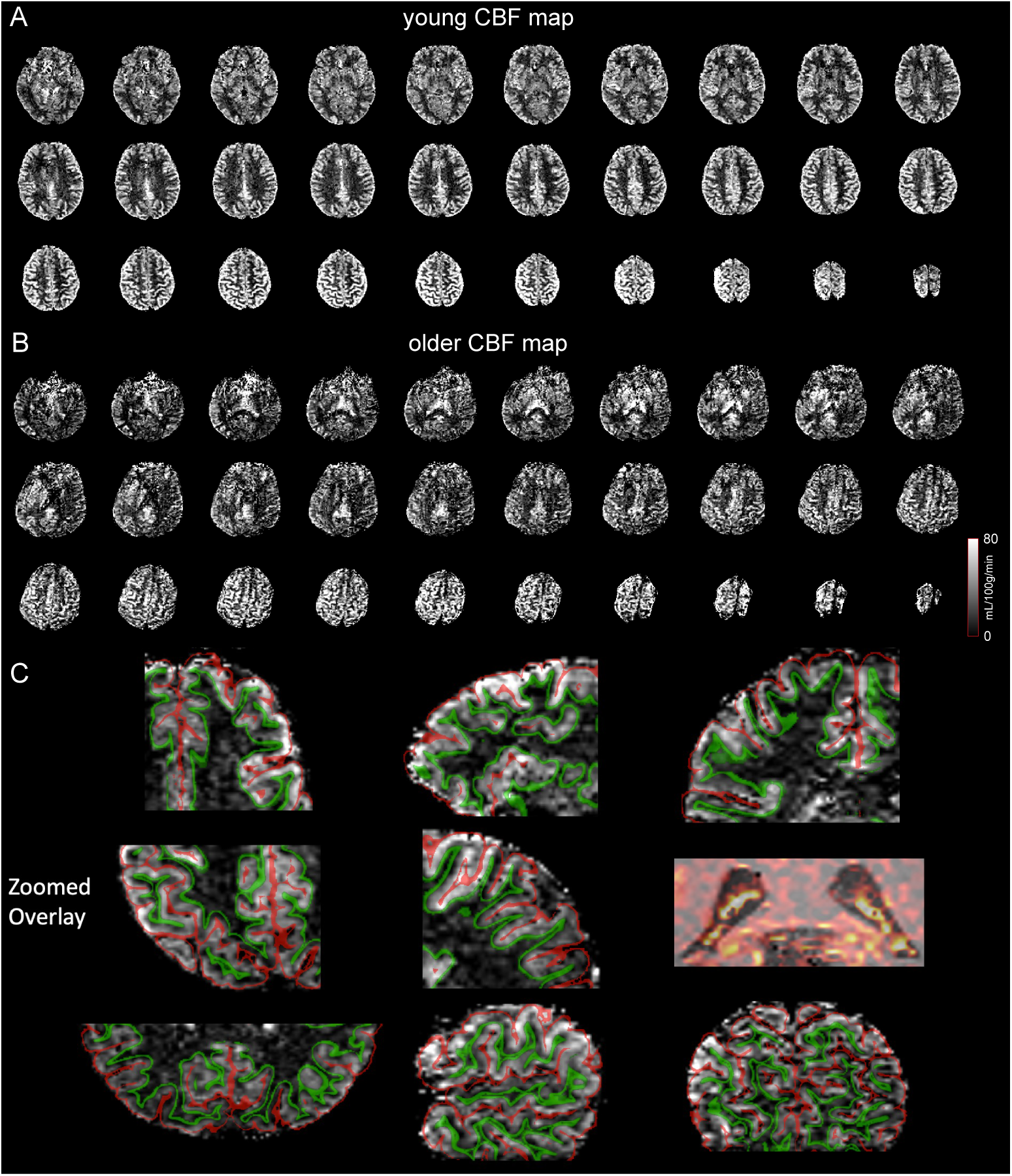
CBF maps show the high-resolution in cortical laminar analysis. (A) CBF map of a young participant. (B) CBF map of an older participant. (C) CBF map zoomed with overlay segmentation. The high image quality and spatial specificity of the CBF map allow for accurate layer-specific analysis and precise display of the choroid plexus with high CBF distribution.

**Figure S3.**
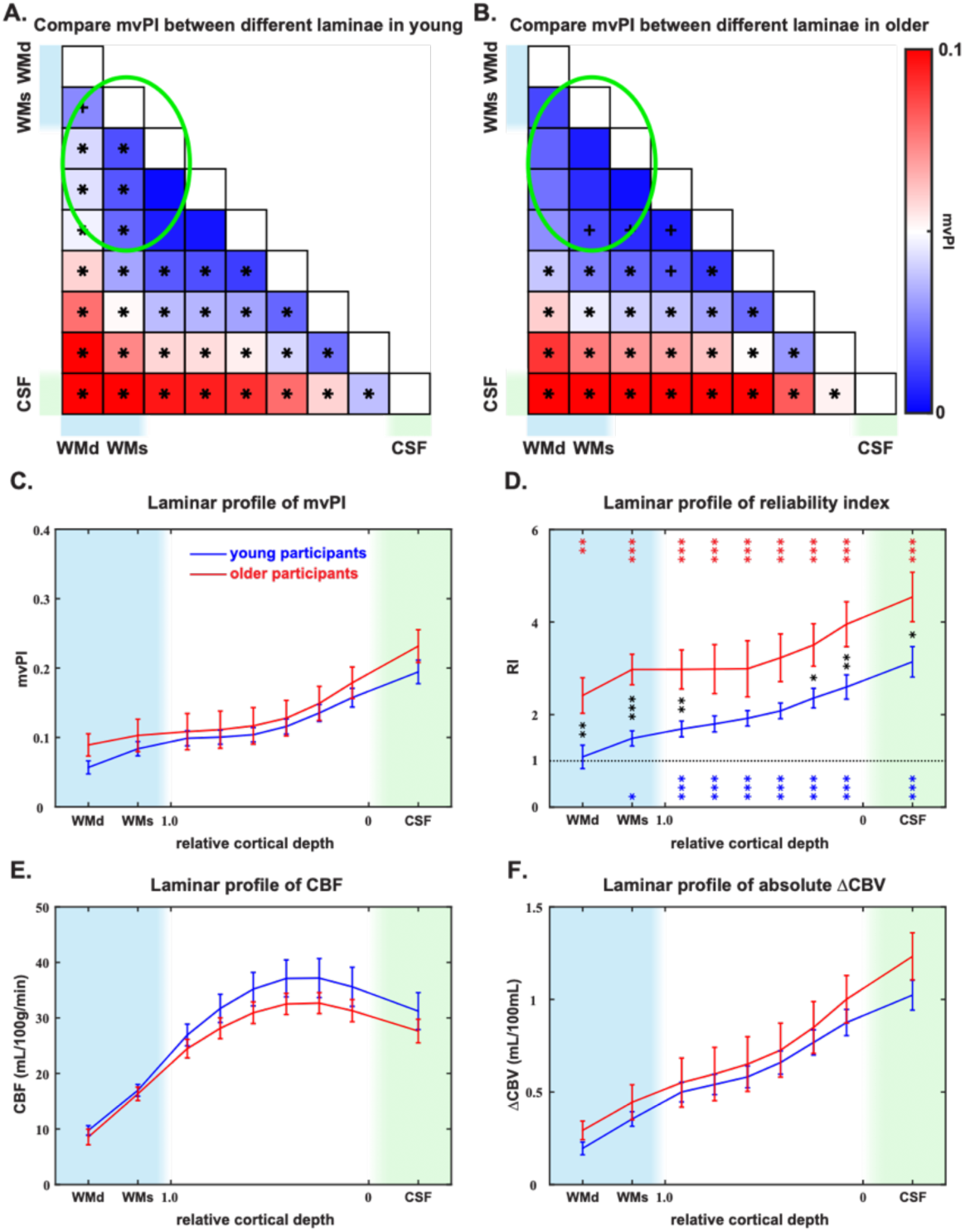
Compare mvPI, RI, CBF and ΔCBV in whole FOV between young and older groups. Matrix of the differences in mvPI values at different depths for young (A) and older (B) participants. Each element of the matrix represents the PI value of the depth corresponding to the row minus the PI value of the depth corresponding to the column. Bonferroni correction, factor = 36; *: corrected p < 0.05; +: uncorrected p < 0.05 but corrected p > 0.05. (C) Laminar profile of mvPI cross young (blue) and older (red) participants. (D) Laminar profile of reliability index (RI) cross young (blue) and older (red) participants. Bonferroni correction, factor = 9; *: corrected p < 0.05; **: corrected p < 0.01; ***: corrected p < 0.001. Red * shows the comparison between older RI and chance-level, blue * shows the comparison between young RI and chance-level, and black * shows the comparison between older RI and young RI. And the laminar profile of CBF (E) and ΔCBV (F) cross young (blue) and older (red) participants.

**Figure S4.**
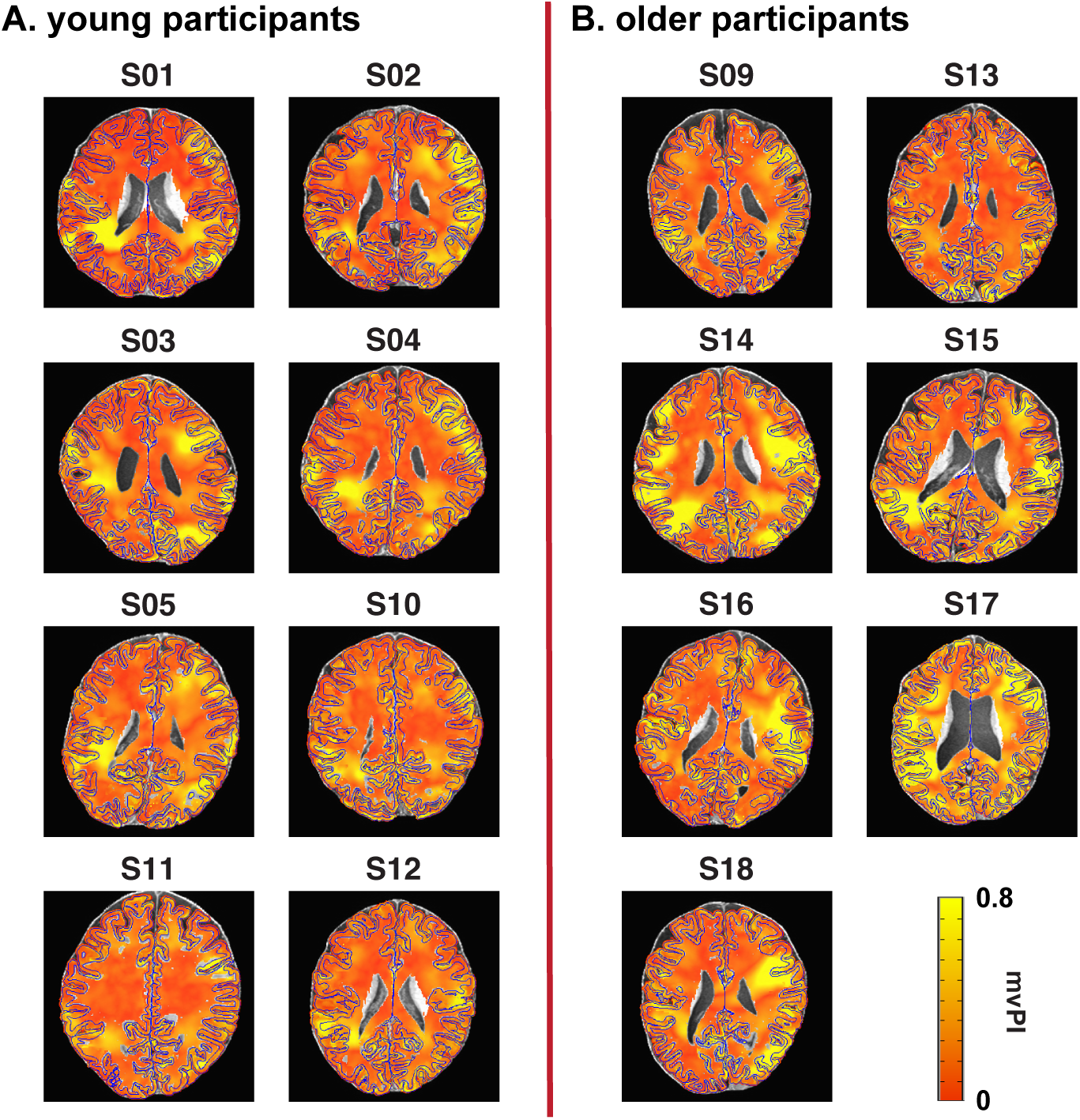
mvPI maps of all other participants. (A) mvPI maps of all other young participants. (B) mvPI maps of all elder participants. As we can see, the mvPI maps of older participants were more variations and were generally higher than these of young participants.

**Figure S5.**
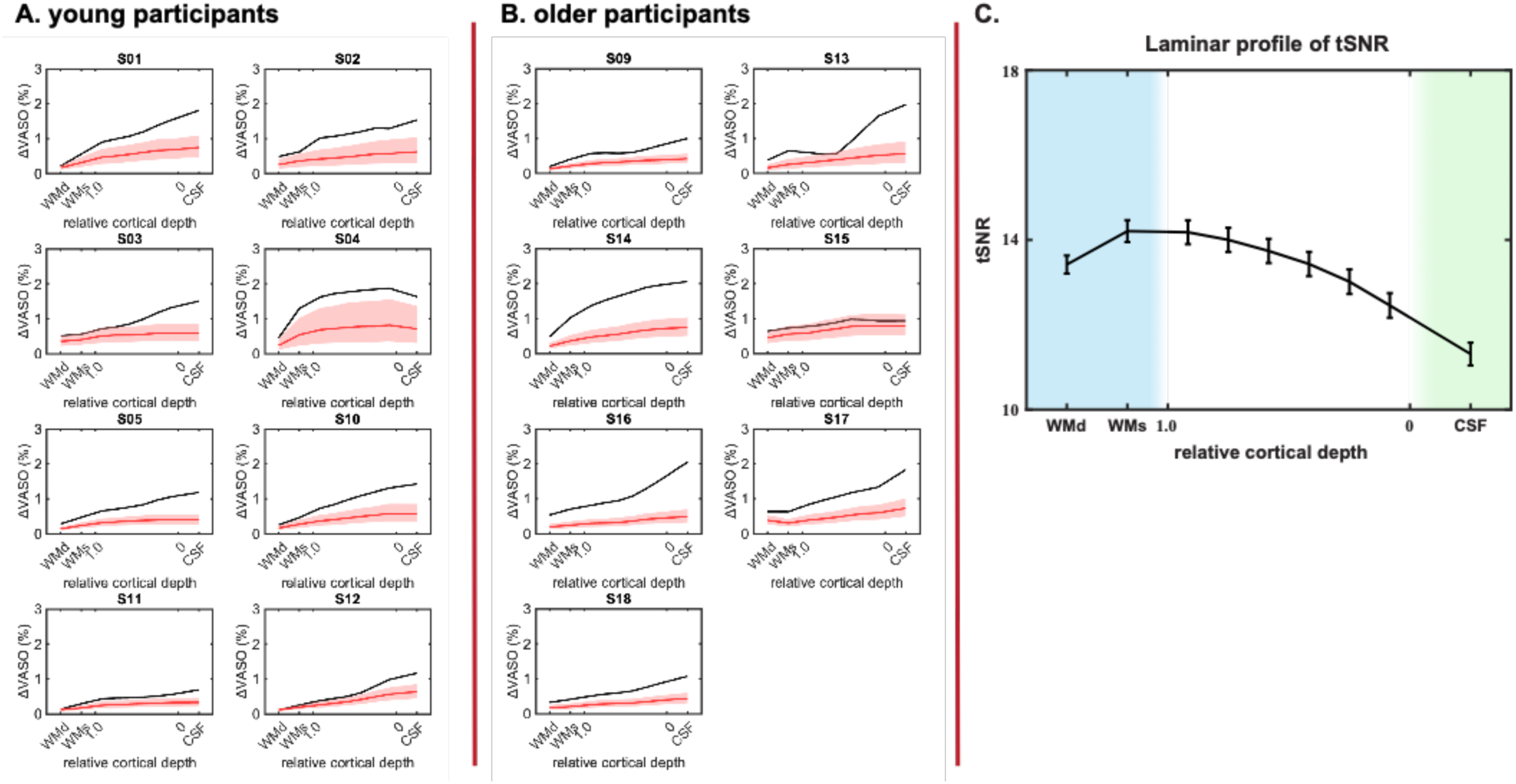
The results of all other participants reliability test. (A) Reliability test results of all other young participants. (B) Reliability test results of all elder participants. As we can see, S15 is an outlier that cannot be distinguished from variations caused by random noise. As a result, the VASO-related group analysis in this study excluded S15 from the final results. And compared with young participants, the ΔVASO of elder were generally significantly higher than chance-level. (C) Laminar tSNR profile. The distribution of tSNR and chance-level are exactly inversely proportional, which illustrates the accuracy of our chance-level calculation.

**Figure S6.**
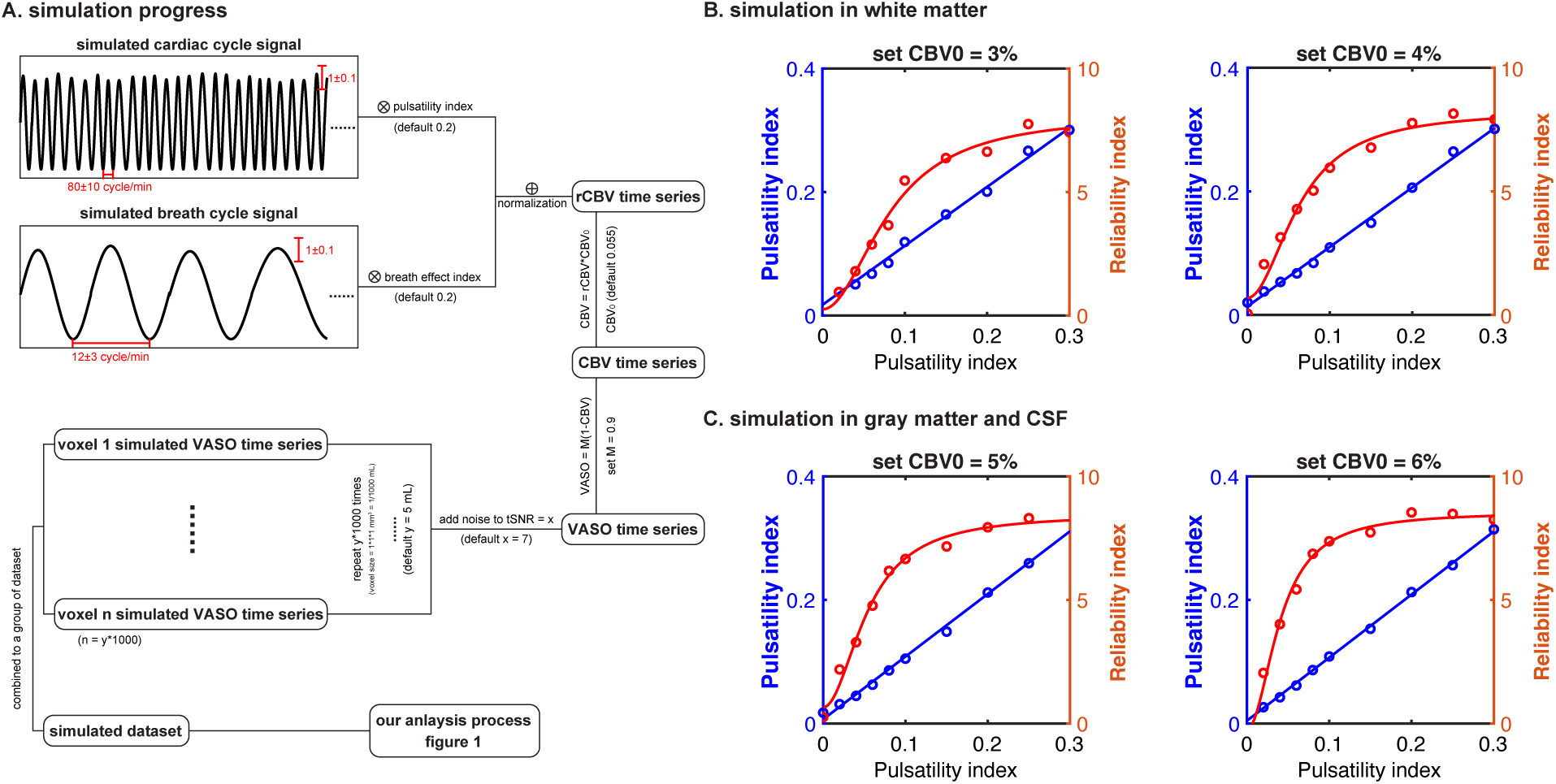
Simulations in white, gray matter and CSF. (A) simulation progress. Details in Method. CBV0 was set to 3%~4% to simulate the conditions in white matter (B), and 5%~6% to simulate the conditions in gray matter and CSF (C), respectively. We found that when CBV0 was high (in gray matter or CSF), the PI value was calculated very accurately and the RI value reached a plateau more quickly. However, when CBV0 was low (in white matter), the vPI value had to be above 0.05 to be accurately calculated. And when the PI value reached 0.1, the calculated RI value could be stable and significantly greater than 1, and it was difficult to reach a plateau. The simulation results explain why we can measure high PI and RI values in the elderly, but they are difficult to detect in young people because the true PI values are smaller.

**Figure S7.**
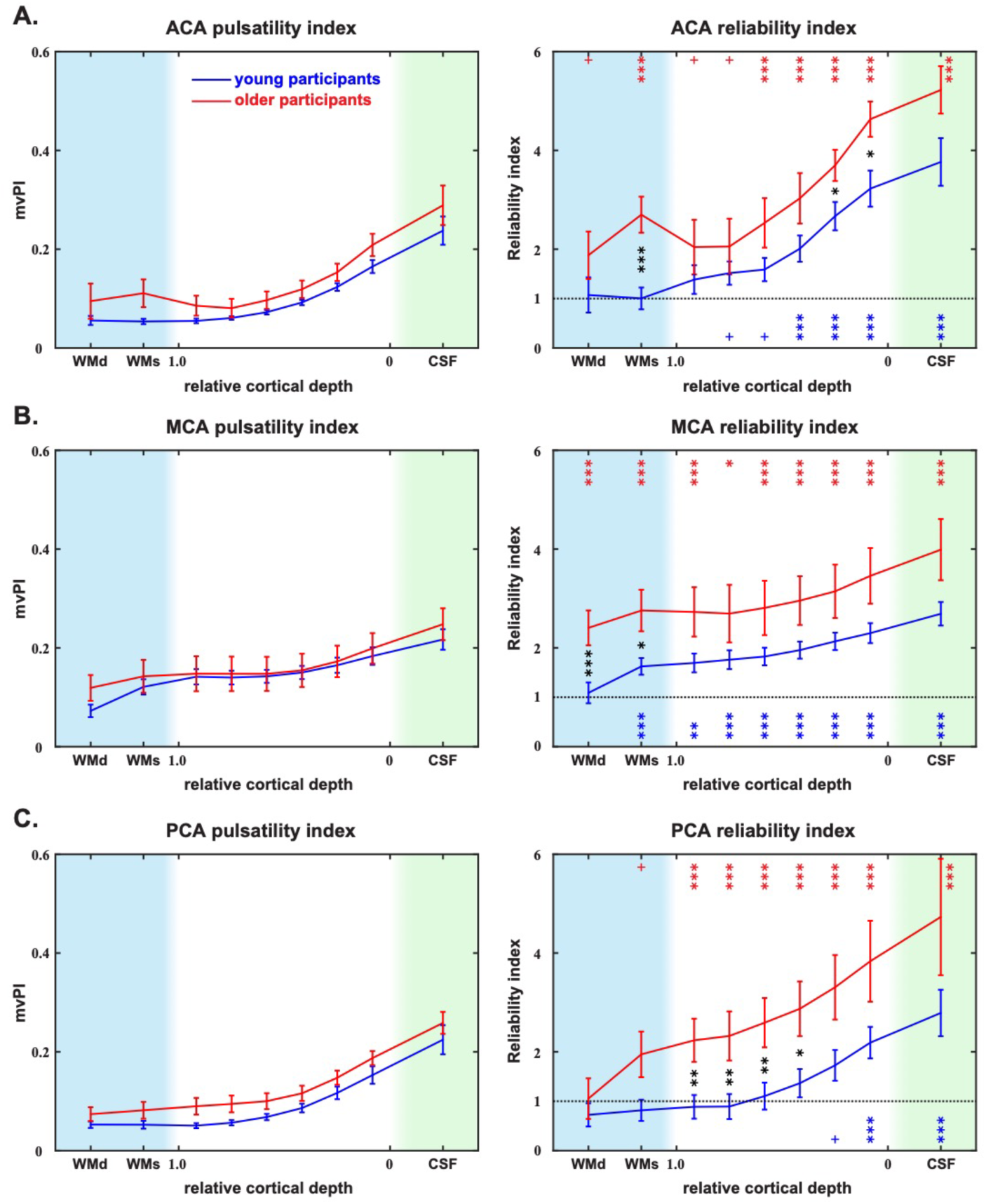
laminar profile of mvPI and RI in ACA, MCA, and PCA. Laminar profiles of mvPI (left) and RI (right) in ACA (A), MCA (B), and PCA (C). The blue and red curves represent the results for younger and elder participants, respectively. The light blue shaded area indicates the white matter area, and the light green shaded area indicates the CSF area. WMd and WMs in x axis represent the deep white matter and superficial white matter. The values on the x-axis from 1 to 0 represent the gray matter from deep to superficial. *: corrected p < 0.05; **: corrected p < 0.01; ***: corrected p < 0.001; +: uncorrected p < 0.05; Error bar is SEM.

**Figure S8.**
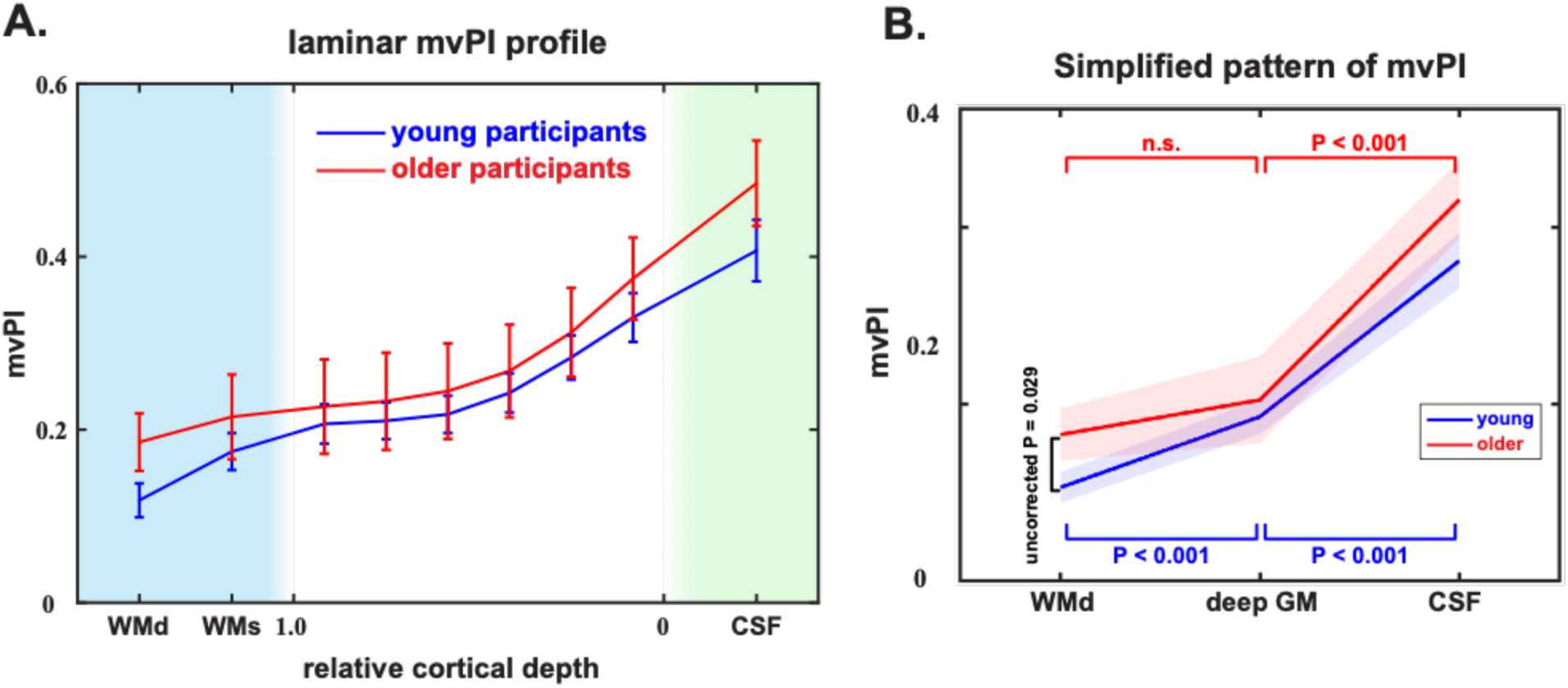
laminar profile of mvPI and simplified pattern using another CBV_0_ value (0.04). (A) Laminar profile of mvPI cross young (blue) and older (red) participants using CBV_0_ = 0.04 (same method with Figure S3C). (B) Simplified pattern of mvPI between young and older (same method with Figure 4C). Compared with the results in the paper, except for the calculated mvPI values, the distribution patterns and statistical results are the same as in the paper. It shows that setting the CBV_0_ value not too different will not have a significant impact on our results.

## REFERENCES

1 Shi, Y., Thrippleton, M. J., Marshall, I. & Wardlaw, J. M. Intracranial pulsatility in patients with cerebral small vessel disease: a systematic review. Clin Sci (Lond) 132, 157–171 (2018).

2 Geurts, L. J., Zwanenburg, J. J. M., Klijn, C. J. M., Luijten, P. R. & Biessels, G. J. Higher Pulsatility in Cerebral Perforating Arteries in Patients With Small Vessel Disease Related Stroke, a 7T MRI Study. Stroke 50, 62–68 (2019).

3 Birnefeld, J., Wahlin, A., Eklund, A. & Malm, J. Cerebral arterial pulsatility is associated with features of small vessel disease in patients with acute stroke and TIA: a 4D flow MRI study. J Neurol 267, 721–730 (2020).

4 Shi, Y. et al. Small vessel disease is associated with altered cerebrovascular pulsatility but not resting cerebral blood flow. J Cereb Blood Flow Metab 40, 85–99 (2020).

5 Lim, J. S., Lee, J. Y., Kwon, H. M. & Lee, Y. S. The correlation between cerebral arterial pulsatility and cognitive dysfunction in Alzheimer’s disease patients. J Neurol Sci 373, 285–288 (2017).

6 Roher, A. E. et al. Transcranial doppler ultrasound blood flow velocity and pulsatility index as systemic indicators for Alzheimer’s disease. Alzheimers Dement 7, 445–455 (2011).

7 Rivera-Rivera, L. A. et al. Changes in intracranial venous blood flow and pulsatility in Alzheimer’s disease: A 4D flow MRI study. J Cereb Blood Flow Metab 37, 2149–2158 (2017).

8 Chung, C. P., Lee, H. Y., Lin, P. C. & Wang, P. N. Cerebral Artery Pulsatility is Associated with Cognitive Impairment and Predicts Dementia in Individuals with Subjective Memory Decline or Mild Cognitive Impairment. J Alzheimers Dis 60, 625–632 (2017).

9 Iliff, J. J. et al. Cerebral arterial pulsation drives paravascular CSF-interstitial fluid exchange in the murine brain. J Neurosci 33, 18190–18199 (2013).

10 Cserr, H. F. & Ostrach, L. H. Bulk flow of interstitial fluid after intracranial injection of blue dextran 2000. Exp Neurol 45, 50–60 (1974).

11 Ichimura, T., Fraser, P. A. & Cserr, H. F. Distribution of extracellular tracers in perivascular spaces of the rat brain. Brain Res 545, 103–113 (1991).

12 Abbott, N. J. Evidence for bulk flow of brain interstitial fluid: significance for physiology and pathology. Neurochem Int 45, 545–552 (2004).

13 Schley, D., Carare-Nnadi, R., Please, C. P., Perry, V. H. & Weller, R. O. Mechanisms to explain the reverse perivascular transport of solutes out of the brain. J Theor Biol 238, 962–974 (2006).

14 Jessen, N. A., Munk, A. S., Lundgaard, I. & Nedergaard, M. The Glymphatic System: A Beginner’s Guide. Neurochem Res 40, 2583–2599 (2015).

15 Engelhardt, B. et al. Vascular, glial, and lymphatic immune gateways of the central nervous system. Acta Neuropathol 132, 317–338 (2016).

16 Louveau, A. et al. Understanding the functions and relationships of the glymphatic system and meningeal lymphatics. J Clin Invest 127, 3210–3219 (2017).

17 Rasmussen, M. K., Mestre, H. & Nedergaard, M. The glymphatic pathway in neurological disorders. Lancet Neurol 17, 1016–1024 (2018).

18 McKnight, C. D., Rouleau, R. M., Donahue, M. J. & Claassen, D. O. The Regulation of Cerebral Spinal Fluid Flow and Its Relevance to the Glymphatic System. Curr Neurol Neurosci Rep 20, 58 (2020).

19 Mestre, H. et al. Flow of cerebrospinal fluid is driven by arterial pulsations and is reduced in hypertension. Nat Commun 9, 4878 (2018).

20 Jammal Salameh, L., Bitzenhofer, S. H., Hanganu-Opatz, I. L., Dutschmann, M. & Egger, V. Blood pressure pulsations modulate central neuronal activity via mechanosensitive ion channels. Science 383, eadk8511 (2024).

21 Thompson, R. S., Trudinger, B. J. & Cook, C. M. Doppler ultrasound waveform indices: A/B ratio, pulsatility index and Pourcelot ratio. Br J Obstet Gynaecol 95, 581–588 (1988).

22 Wahlin, A. et al. Measuring pulsatile flow in cerebral arteries using 4D phase-contrast MR imaging. AJNR Am J Neuroradiol 34, 1740–1745 (2013).

23 Wahlin, A. et al. Assessment of craniospinal pressure-volume indices. AJNR Am J Neuroradiol 31, 1645–1650 (2010).

24 van den Kerkhof, M., Jansen, J. F. A., van Oostenbrugge, R. J. & Backes, W. H. 1D versus 3D blood flow velocity and pulsatility measurements of lenticulostriate arteries at 7T MRI. Magn Reson Imaging 96, 144–150 (2023).

25 Huber, L. et al. High-Resolution CBV-fMRI Allows Mapping of Laminar Activity and Connectivity of Cortical Input and Output in Human M1. Neuron 96, 1253–1263 e1257 (2017).

26 Zhao, C. et al. Whole-Cerebrum distortion-free three-dimensional pseudo-continuous arterial spin labeling at 7T. Neuroimage 277, 120251 (2023).

27 Duvernoy, H. M., Delon, S. & Vannson, J. L. Cortical blood vessels of the human brain. Brain Res Bull 7, 519–579 (1981).

28 Akbari, A., Bollmann, S., Ali, T. S. & Barth, M. Modelling the depth-dependent VASO and BOLD responses in human primary visual cortex. Hum Brain Mapp 44, 710–726 (2023).

29 Warnert, E. A., Murphy, K., Hall, J. E. & Wise, R. G. Noninvasive assessment of arterial compliance of human cerebral arteries with short inversion time arterial spin labeling. J Cereb Blood Flow Metab 35, 461–468 (2015).

30 Kim, D. E. et al. Mapping the Supratentorial Cerebral Arterial Territories Using 1160 Large Artery Infarcts. JAMA Neurol 76, 72–80 (2019).

31 Schirmer, M. D. et al. Spatial Signature of White Matter Hyperintensities in Stroke Patients. Front Neurol 10, 208 (2019).

32 Farkas, E. & Luiten, P. G. Cerebral microvascular pathology in aging and Alzheimer’s disease. Prog Neurobiol 64, 575–611 (2001).

33 Kelly, R., Hayward, C., Avolio, A. & O’Rourke, M. Noninvasive determination of age-related changes in the human arterial pulse. Circulation 80, 1652–1659 (1989).

34 Garrett, D. D., Lindenberger, U., Hoge, R. D. & Gauthier, C. J. Age differences in brain signal variability are robust to multiple vascular controls. Sci Rep 7, 10149 (2017).

35 Kannurpatti, S. S., Motes, M. A., Rypma, B. & Biswal, B. B. Neural and vascular variability and the fMRI-BOLD response in normal aging. Magn Reson Imaging 28, 466–476 (2010).

36 Pahlavian, S. H. et al. Cerebroarterial pulsatility and resistivity indices are associated with cognitive impairment and white matter hyperintensity in elderly subjects: A phase-contrast MRI study. J Cereb Blood Flow Metab 41, 670–683 (2021).

37 van Hespen, K. M., Kuijf, H. J., Hendrikse, J., Luijten, P. R. & Zwanenburg, J. J. M. Blood Flow Velocity Pulsatility and Arterial Diameter Pulsatility Measurements of the Intracranial Arteries Using 4D PC-MRI. Neuroinformatics 20, 317–326 (2022).

38 van Tuijl, R. J. et al. Does the Internal Carotid Artery Attenuate Blood-Flow Pulsatility in Small Vessel Disease? A 7 T 4D-Flow MRI Study. J Magn Reson Imaging 56, 527–535 (2022).

39 Henry-Feugeas, M. C. & Koskas, P. Cerebral vascular aging: extending the concept of pulse wave encephalopathy through capillaries to the cerebral veins. Curr Aging Sci 5, 157–167 (2012).

40 Tang, J. et al. Assessment of arterial pulsatility of cerebral perforating arteries using 7T high-resolution dual-VENC phase-contrast MRI. Magn Reson Med (2024).

41 Li, Y. et al. Three-dimensional assessment of brain arterial compliance: Technical development, comparison with aortic pulse wave velocity, and age effect. Magn Reson Med 86, 1917–1928 (2021).

42 Richard, K. Cardiovascular physiology concepts. Lippincott Williams & Wilkins (2011).

43 Faraci, F. M. & Heistad, D. D. Regulation of large cerebral arteries and cerebral microvascular pressure. Circ Res 66, 8–17 (1990).

44 Davis, M. J. & Hill, M. A. Signaling mechanisms underlying the vascular myogenic response. Physiol Rev 79, 387–423 (1999).

45 Lacolley, P., Regnault, V., Nicoletti, A., Li, Z. & Michel, J. B. The vascular smooth muscle cell in arterial pathology: a cell that can take on multiple roles. Cardiovasc Res 95, 194–204 (2012).

46 Safar, M. E., Levy, B. I. & Struijker-Boudier, H. Current perspectives on arterial stiffness and pulse pressure in hypertension and cardiovascular diseases. Circulation 107, 2864–2869 (2003).

47 Weller, R. O., Boche, D. & Nicoll, J. A. Microvasculature changes and cerebral amyloid angiopathy in Alzheimer’s disease and their potential impact on therapy. Acta Neuropathol 118, 87–102 (2009).

48 Yousif, L. F., Di Russo, J. & Sorokin, L. Laminin isoforms in endothelial and perivascular basement membranes. Cell Adh Migr 7, 101–110 (2013).

49 Iadecola, C. Neurovascular regulation in the normal brain and in Alzheimer’s disease. Nat Rev Neurosci 5, 347–360 (2004).

50 J, C. M. The cerebral circulation. (2016).

51 Iliff, J. J. et al. A paravascular pathway facilitates CSF flow through the brain parenchyma and the clearance of interstitial solutes, including amyloid beta. Sci Transl Med 4, 147ra111 (2012).

52 Iliff, J. J. et al. Brain-wide pathway for waste clearance captured by contrast-enhanced MRI. J Clin Invest 123, 1299–1309 (2013).

53 Cserr, H. F., Cooper, D. N., Suri, P. K. & Patlak, C. S. Efflux of radiolabeled polyethylene glycols and albumin from rat brain. Am J Physiol 240, F319–328 (1981).

54 Rennels, M. L., Gregory, T. F., Blaumanis, O. R., Fujimoto, K. & Grady, P. A. Evidence for a ‘paravascular’ fluid circulation in the mammalian central nervous system, provided by the rapid distribution of tracer protein throughout the brain from the subarachnoid space. Brain Res 326, 47–63 (1985).

55 Rennels, M. L., Blaumanis, O. R. & Grady, P. A. Rapid solute transport throughout the brain via paravascular fluid pathways. Adv Neurol 52, 431–439 (1990).

56 Hajdu, M. A., McElmurry, R. T., Heistad, D. D. & Baumbach, G. L. Effects of aging on cerebral vascular responses to serotonin in rats. Am J Physiol 264, H2136–2140 (1993).

57 Jiang, H. X., Chen, P. C., Sobin, S. S. & Giannotta, S. L. Age related alterations in the response of the pial arterioles to adenosine in the rat. Mech Ageing Dev 65, 257–276 (1992).

58 Tarumi, T. et al. Cerebral hemodynamics in normal aging: central artery stiffness, wave reflection, and pressure pulsatility. J Cereb Blood Flow Metab 34, 971–978 (2014).

59 Linville, D. G. & Arneric, S. P. Cortical cerebral blood flow governed by the basal forebrain: age-related impairments. Neurobiol Aging 12, 503–510 (1991).

60 Lacombe, P., Sercombe, R., Vaucher, E. & Seylaz, J. Reduced cortical vasodilatory response to stimulation of the nucleus basalis of Meynert in the aged rat and evidence for a control of the cerebral circulation. Ann N Y Acad Sci 826, 410–415 (1997).

61 Abernethy, W. B., Bell, M. A., Morris, M. & Moody, D. M. Microvascular density of the human paraventricular nucleus decreases with aging but not hypertension. Exp Neurol 121, 270–274 (1993).

62 Amenta, F. et al. Age-related changes in brain microanatomy: sensitivity to treatment with the dihydropyridine calcium channel blocker darodipine (PY 108-068). Brain Res Bull 36, 453–460 (1995).

63 Amenta, F. et al. Effect of long-term treatment with the dihydropyridine-type calcium channel blocker darodipine (PY 108-068) on the cerebral capillary network in aged rats. Mech Ageing Dev 78, 27–37 (1995).

64 Sonntag, W. E., Lynch, C. D., Cooney, P. T. & Hutchins, P. M. Decreases in cerebral microvasculature with age are associated with the decline in growth hormone and insulin-like growth factor 1. Endocrinology 138, 3515–3520 (1997).

65 de Jong, G. I., de Weerd, H., Schuurman, T., Traber, J. & Luiten, P. G. Microvascular changes in aged rat forebrain. Effects of chronic nimodipine treatment. Neurobiol Aging 11, 381–389 (1990).

66 de Jong, G. I., Jansen, A. S., Horvath, E., Gispen, W. H. & Luiten, P. G. Nimodipine effects on cerebral microvessels and sciatic nerve in aging rats. Neurobiol Aging 13, 73–81 (1992).

67 Luiten, P. G., de Jong, G. I. & Schuurman, T. Cerebrovascular, neuronal, and behavioral effects of long-term Ca2+ channel blockade in aging normotensive and hypertensive rat strains. Ann N Y Acad Sci 747, 431–451 (1994).

68 Keuker, J. I., Luiten, P. G. & Fuchs, E. Capillary changes in hippocampal CA1 and CA3 areas of the aging rhesus monkey. Acta Neuropathol 100, 665–672 (2000).

69 Sun, Z. Aging, arterial stiffness, and hypertension. Hypertension 65, 252–256 (2015).

70 Mitchell, G. F. et al. Changes in arterial stiffness and wave reflection with advancing age in healthy men and women: the Framingham Heart Study. Hypertension 43, 1239–1245 (2004).

71 Gorelick, P. B. et al. Vascular contributions to cognitive impairment and dementia: a statement for healthcare professionals from the american heart association/american stroke association. Stroke 42, 2672–2713 (2011).

72 Prins, N. D. et al. Cerebral white matter lesions and the risk of dementia. Arch Neurol 61, 1531–1534 (2004).

73 Pantoni, L. Cerebral small vessel disease: from pathogenesis and clinical characteristics to therapeutic challenges. Lancet Neurol 9, 689–701 (2010).

74 Wardlaw, J. M., Smith, C. & Dichgans, M. Small vessel disease: mechanisms and clinical implications. Lancet Neurol 18, 684–696 (2019).

75 Knopman, D. S. et al. Alzheimer disease. Nat Rev Dis Primers 7, 33 (2021).

76 Purkayastha, S. et al. Impaired cerebrovascular hemodynamics are associated with cerebral white matter damage. J Cereb Blood Flow Metab 34, 228–234 (2014).

77 Coenen, M. et al. Spatial distributions of white matter hyperintensities on brain MRI: A pooled analysis of individual participant data from 11 memory clinic cohorts. Neuroimage Clin 40, 103547 (2023).

78 Talagala, S. L., Sarlls, J. E., Liu, S. & Inati, S. J. Improvement of temporal signal-to-noise ratio of GRAPPA accelerated echo planar imaging using a FLASH based calibration scan. Magn Reson Med 75, 2362–2371 (2016).

79 Huber, L., Uludag, K. & Moller, H. E. Non-BOLD contrast for laminar fMRI in humans: CBF, CBV, and CMRO2. Neuroimage 197, 742–760 (2019).

80 Stirnberg, R. & Stocker, T. Segmented K-space blipped-controlled aliasing in parallel imaging for high spatiotemporal resolution EPI. Magn Reson Med 85, 1540–1551 (2021).

81 Zhao C. et al. Iso-1.25mm Whole-cerebrum pCASL at 7T for Mapping Depth-dependent Cortical Gray Matter and Tract-specific White Matter Cerebral Blood Flow. ISMRM 1264 (2024).

82 Cox, R. W. AFNI: software for analysis and visualization of functional magnetic resonance neuroimages. Comput Biomed Res 29, 162–173 (1996).

83 Dale, A. M., Fischl, B. & Sereno, M. I. Cortical surface-based analysis. I. Segmentation and surface reconstruction. Neuroimage 9, 179–194 (1999).

84 Avants, B. B. et al. A reproducible evaluation of ANTs similarity metric performance in brain image registration. Neuroimage 54, 2033–2044 (2011).

85 Friston, K. J. et al. Statistical parametric maps in functional imaging: A general linear approach. Human Brain Mapping 2, 189–210 (1995).

86 Saad, Z. S. et al. A new method for improving functional-to-structural MRI alignment using local Pearson correlation. Neuroimage 44, 839–848 (2009).

87 Waehnert, M. D. et al. Anatomically motivated modeling of cortical laminae. Neuroimage 93 Pt 2, 210–220 (2014).

88 van den Kerkhof, M., et al. Impaired damping of cerebral blood flow velocity pulsatility is associated with the number of perivascular spaces as measured with 7T MRI. J Cereb Blood Flow Metab 43, 937–946 (2023).

89 Lu, H., Golay, X., Pekar, J. J. & Van Zijl, P. C. Functional magnetic resonance imaging based on changes in vascular space occupancy. Magn Reson Med 50, 263–274 (2003).

90 Grubb, R. L., Jr., Raichle, M. E., Eichling, J. O. & Ter-Pogossian, M. M. The effects of changes in PaCO2 on cerebral blood volume, blood flow, and vascular mean transit time. Stroke 5, 630–639 (1974).

91 Lin, A. L. et al. Evaluation of MRI models in the measurement of CMRO2 and its relationship with CBF. Magn Reson Med 60, 380–389 (2008).

92 Liu, C. et al. Layer-dependent multiplicative effects of spatial attention on contrast responses in human early visual cortex. Prog Neurobiol 207, 101897 (2021).

93 Shahzad Ahmad Qureshi, Sikander M. Mirza & Arif, M. Fitness Function Evaluation for Image Reconstruction using Binary Genetic Algorithm for Parallel Ray Transmission Tomography. 2006 2nd International Conference on Emerging Technologies. IEEE 7, 196–201 (2006).

94 Hore, A. & Ziou, D. Image Quality Metrics: PSNR vs. SSIM. 2010 20th International Conference on Pattern Recognition, 2366–2369 (2010).

95 M, K. J. An introduction into autonomic nervous function. Physiological measurement 38, R89 (2017).

96 Glover, G. H., Li, T. Q. & Ress, D. Image-based method for retrospective correction of physiological motion effects in fMRI: RETROICOR. Magn Reson Med 44, 162–167 (2000).

97 van der Kleij, L. A., De Vis, J. B., de Bresser, J., Hendrikse, J. & Siero, J. C. W. Arterial CO(2) pressure changes during hypercapnia are associated with changes in brain parenchymal volume. Eur Radiol Exp 4, 17 (2020).

